# Rare coding variation implicates thirteen genes in bipolar disorder across 232,536 individuals from global populations

**DOI:** 10.64898/2026.09.30.26364416

**Authors:** Calwing Liao, Robert Ye, Julia M. Sealock, Toni Boltz, Franjo Ivankovic, Hilary Finucane, Daniel Howrigan, Yijia Christiana Liu, F. Kyle Satterstrom, Arsalan Hassan, Melkam Alemayehu, Stella Gichuru, Rehema M. Mwende, Charles R.J.C. Newton, Nastassja Koen, Zukiswa Zingela, Ana M. Diaz-Zuluaga, Ana M. Ramirez-Diaz, Victor I. Reus, Terri Teshiba, Aarno Palotie, Eija Hämäläinen, Olli Pietiläinen, Penelope A. Lind, Dan J. Siskind, Ian B. Hickie, Pamela Morales-Cedillo, Joanna Jiménez-Pavón, Marco Antonio Sanabrais-Jiménez, Carlo Esteban Sotelo-Ramírez, Eric Hahn, Van Phi Nguyen, Elizabeth Karlson, Chiao-Erh Chang, Hsi-Chung Chen, Martin Alda, Mikael Landén, Jordan W. Smoller, Nicholas Craddock, Marquis P. Vawter, David Curtis, Andrew McQuillin, René S. Kahn, Roel A. Ophoff, Annabel Vreeker, Christina Hultman, Patrick F. Sullivan, Michael E. Talkowski, Douglas H. Blackwood, Andrew McIntosh, Ann E. Pulver, Bruce Cohen, Rolf Adolfsson, Andreas Reif, Fernando Goes, Robert Yolken, Aiden P. Corvin, Derek W. Morris, BIPEX Collection Scientists, Felecia Cerrato, Sinéad B. Chapman, Caroline Cusick, Zhenglin Guo, Ana Maria Olivares, Guy A. Rouleau, Biju Viswanath, Po-Hsiu Kuo, Van Tuan Nguyen, Thi Minh Tam Ta, Beatriz Camarena, Carlos N. Pato, Michele T. Pato, Sarah E. Medland, Nelson Freimer, Loes Olde Loohuis, Carlos Lopez-Jaramillo, Rocky Stroud, Lukoye Atwoli, Akena Dickens, Karestan C. Koenen, Symon M. Kariuki, Solomon Teferra, Dan J. Stein, Muhammad Ayub, James A. Knowles, Mark J. Daly, Hailiang Huang, Benjamin M. Neale

## Abstract

Bipolar disorder (BD) is highly heritable, yet the contribution of rare coding variation remains incompletely characterized. We analyzed sequencing data from 64,435 individuals with BD and 168,101 controls spanning multiple ancestries and 22 countries, representing the largest and most global sequencing resource with a 6.7-fold increase in effective sample size over the previous study iteration. We observe enrichment of protein-truncating and damaging missense variants in constrained genes and curated neuropsychiatric gene sets, with no enrichment of synonymous variation. These enrichment signals were consistent across ancestry groups, suggesting that genetic risk factors for BD are consistent worldwide. Gene-level analyses identified 13 exome-wide significant genes and an additional 20 genes at FDR < 0.05. These genes showed convergence with common and rare variant risk across other neuropsychiatric disorders. Expression analyses also showed preferential brain expression and increased developmental expression during early childhood. Modelling 3D protein structures further highlighted clustering of ultra-rare missense variants at a predicted interaction interfaces in two Bonferroni-significant genes, *DOP1A* and *ATP9A*; a potential mechanism linking membrane trafficking to BD risk. Together, these results implicate a constellation of rare variants that map onto neuronal biology and demonstrate that diverse global populations converge on shared genetic signals underlying BD risk.

## Introduction

Bipolar disorder (BD) affects approximately 1–2% of the global population and is characterized by recurrent episodes of mania and depression that contribute to excess morbidity, mortality, and socioeconomic burden^1^. Twins and family studies estimate the heritability of BD from 60% to 85%^2,3^. Genome-wide association studies (GWAS) have identified hundreds of common variants of small effect^4,5^. In contrast, the contribution of rare coding variants, an important source of high-impact variation in several neurodevelopmental and psychiatric disorders, has remained less clearly defined for BD.

The first Bipolar Exome (BipEx) meta-analysis focused on European ancestry individuals comprising of 13,933 cases and 14,422 controls, and demonstrated enrichment of ultra-rare protein-truncating variants (PTV) in constrained genes but lacked sufficient power to discover individual genes at exome-wide significance^6^. Combining BipEx data with schizophrenia (SCZ) sequencing studies identified *AKAP11* as a definitive shared risk gene. A follow-up study incorporating additional BD cases to BipEx 1.0 strengthened evidence for *AKAP11*, *HECTD2* and suggested additional candidate genes^7^.

Defining this architecture requires substantially larger cohorts^8^. Given that ultra rare variants are individually infrequent, even variants with relatively large effects may be observed in only a small number of individuals, limiting statistical power for gene discovery and precise estimation of genetic effects. Continued expansion of sequencing cohorts will therefore be essential to identify additional risk genes, characterize the spectrum of variation within them, and refine our understanding of the rare-variant contribution to psychiatric disorders. Increasing representation across populations will further improve the breadth and generalizability of these findings^9^.

Here, we present an expanded BipEx 2.0 resource and analysis comprising 64,435 individuals with BD and 168,101 controls assembled from global cohorts spanning multiple continental ancestries. Using joint variant calling and uniform quality control, we systematically evaluate the contribution of ultra-rare coding variation to BD risk. We test for enrichment of PTVs and damaging missense variants, identify associated genes, and assess the biological coherence of these signals through analyses of gene constraint, neuropsychiatric gene sets, common-variant heritability enrichment, and brain expression across tissues, cell types, and developmental stages. We further examine structural clustering of missense variants within predicted three-dimensional protein structures to generate mechanistic hypotheses for individual BD risk genes. Together, these analyses expand and refine the contribution of rare coding variation to BD risk. Integrating global cohorts of large-scale sequencing data with functional and 3D protein information reveals a constellation of rare and common genetic variation that converges on neuronal biology and represents substantial discovery in BD biology.

## Results

### Cohorts, sequencing, and variant annotation

We analyzed 88 cohorts spanning five [AFR, AMR, EAS, EUR, SAS] continental ancestries (**Figure 1**) across 22 countries. Subsequently, we applied standardized sample-level QC (sex check, DNA-level contamination, sample duplicates/relatives, sequencing coverage outliers, ancestry inference) and variant-level QC (genotype quality, call rate, depth, allele balance, sequencing platform harmonization) detailed in the Supplementary Methods. After QC, 232,536 individuals (64,435 cases; 168,101 controls) remained with 48,720,030 variants. Within each global ancestry group, we had 105,673 EUR, 38,702 AFR, 52,641 AMR, 17,987 EAS, 17,533 SAS. This represents an additional 50,502 BD cases and 153,679 controls relative to BipEx 1.0. Variant level functional annotation used Variant Effect Predictor (VEP) v95, and we only included variants with protein-coding consequences. PTVs included stop-gain, frameshift, transcript ablation, splice donor and splice acceptor categories^10^. To integrate missense pathogenicity evidence, we computed a composite mean-rank across MPC, AlphaMissense, and MisFitS for each variant and defined “damaging missense” at ≥96th percentile of the composite distribution unless otherwise stated^11–13^. Gene-level constraint was indexed by GISMO-mis and sHet^14,15^ (**Supplementary Figure 9**).

**Figure 1.**
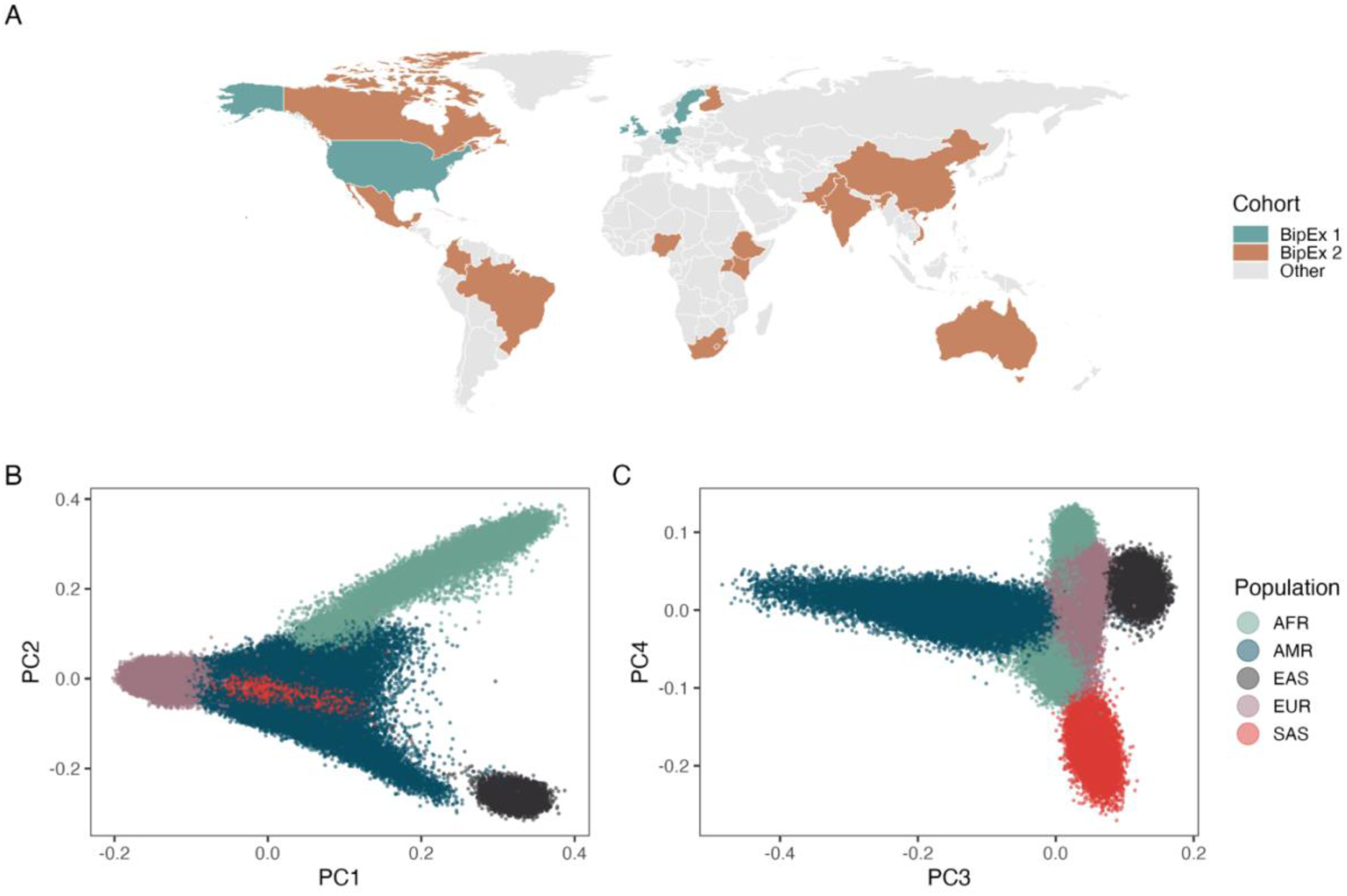
Global cohort distribution and genetic ancestry structure of BipEx samples. **(A)** Geographic distribution of participating cohorts included in BipEx 1 and BipEx 2. Countries contributing samples to BipEx 1 and BipEx 2 are highlighted in teal and rust, respectively; all other countries are shown in grey. **(B)** Principal component analysis (PC1 vs PC2) of BipEx samples passing QC, colored by inferred continental ancestry. Each point represents an individual. **(C)** Principal component analysis (PC3 vs PC4) of the same individuals, colored by inferred continental ancestry. AFR = African; AMR = Admixed American; EAS = East Asian; EUR = European; SAS = South Asian.

### Gene-set burden analyses implicate constrained and neuropsychiatric genes in bipolar disorder

Gene-set burden analyses in BD cases identified robust enrichment of rare deleterious variation across multiple neuropsychiatric gene sets. PTVs were significantly enriched among cases within constrained genes (OR 1.12, 95% CI 1.09–1.16, P = 9.9 × 10^-14^), as well as curated Autism Spectrum Disorder (ASD), SCZ, and neurodevelopmental disorders (NDD) gene sets (ASD OR 1.24, P = 1.8 × 10^-10^; SCZ OR 1.93, P = 9.9 × 10^-22^; NDD OR 1.12, P = 8.8 × 10^-6^) (**Figure 2a**)^16–18^. Damaging missense variants showed a similar pattern of enrichment, with significant effects observed for constrained genes (OR 1.13, P = 2.4 × 10^-20^), ASD (OR 1.21, P = 7.6 × 10^-10^), and NDD gene sets (OR 1.19, P = 3.4 × 10^-12^). Enrichment for SCZ genes under the damaging missense model was even stronger (OR 1.40, P = 1.8 × 10^-3^).

**Figure 2.**
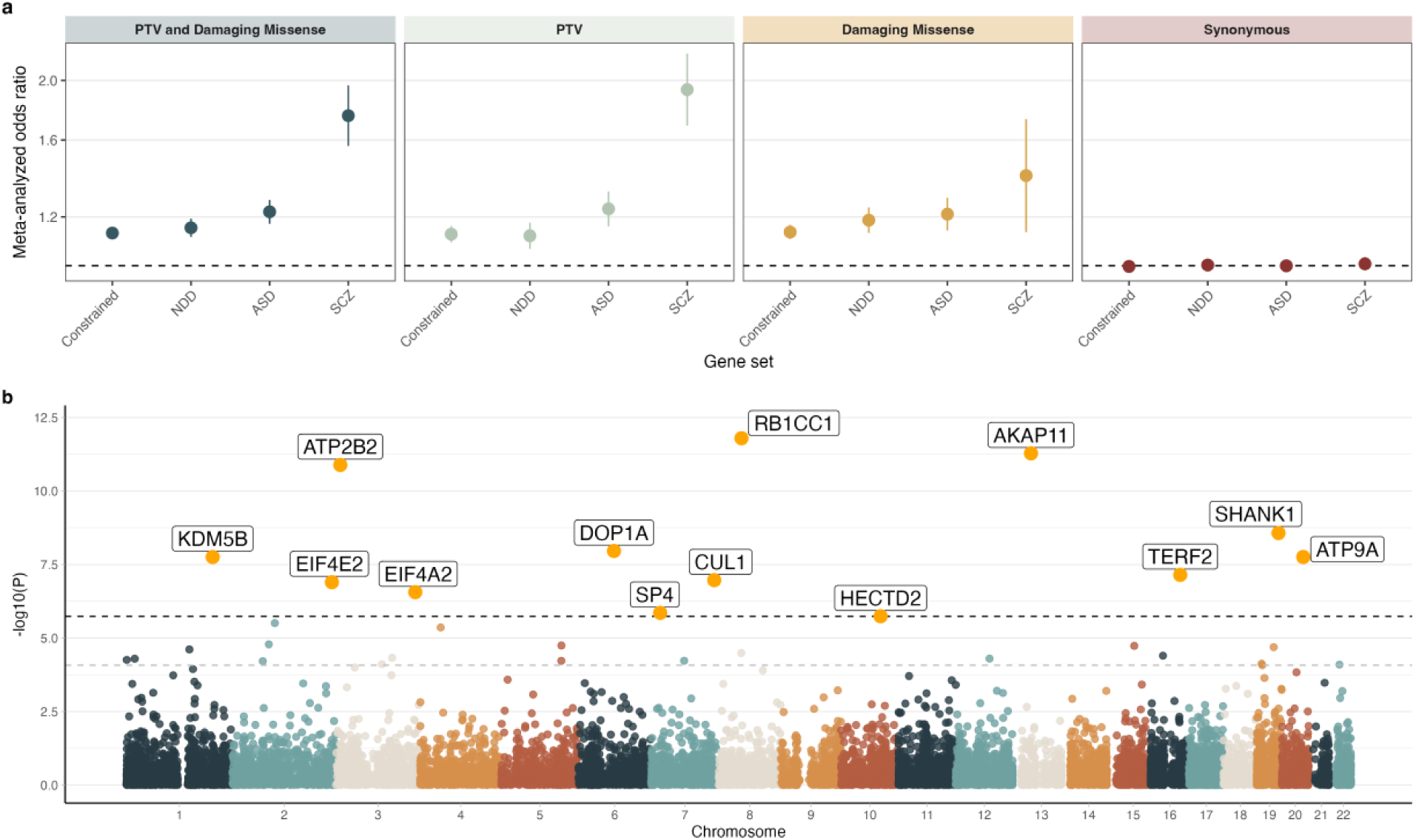
Rare variant burden across gene sets and gene-level association in bipolar disorder. **(a)** Meta-analyzed odds ratios for rare variant burden across predefined gene sets, stratified by variant class. Results are shown for PTVs and damaging missense variants combined (PTV + damaging missense), PTVs, damaging missense variants, and synonymous variants. Points denote meta-analyzed odds ratios, and vertical lines indicate 95% confidence intervals. The dashed horizontal line indicates an odds ratio of 1. Gene sets include constrained genes (n = 3570), neurodevelopmental disorders (NDD) genes (n = 373), ASD genes (n = 72), and SCZ genes (n = 32). **(b)** Gene-level Manhattan plot of rare variant association results across the genome. Each point represents a gene, plotted by chromosomal position and −log10(P-value). Genes exceeding the significance threshold are highlighted in orange and labeled. The darker dashed horizontal line indicates the Bonferroni exome-wide significance threshold, and the lighter grey indicates the 5% FDR significance threshold.

When protein truncating and damaging missense variants were combined, effect sizes were similarly consistently increased across all gene sets, with the strongest enrichment observed for SCZ genes (OR 1.75, 95% CI 1.56–1.96, P = 2.7 × 10^-22^), followed by ASD (OR 1.22, P = 1.1 × 10^-18^), NDD (OR 1.15, P = 6.3 × 10^-16^), and constrained genes (OR 1.13, P = 3.5 × 10^-32^). In contrast, synonymous variants showed no evidence of enrichment for any gene set (all OR ≈ 1.00, all P > 0.05), serving as a negative control.

We found enrichment signals were largely consistent across ancestry strata. Although moderate heterogeneity was observed for several gene sets, particularly for PTVs (I^2^ range 57–75%, **Supplementary Table 4**), the direction of effect were broadly uniform across ancestries (**Supplementary Table 3**), and fixed-effect inverse-variance weighted meta-analysis yielded highly significant associations (**Supplementary Table 4**). Together, these results demonstrate convergent enrichment of rare loss-of-function (LoF) and missense variants in biologically constrained and neurodevelopmentally relevant gene sets among individuals with BD across diverse populations.

### Gene-level associations identify 13 exome-wide significant BD genes

We performed gene-level ultra-rare variant association testing using a Cochran-Mantel-Haenszel (CMH) framework, testing ultra-rare PTVs and damaging missense variants within genes stratified across 24 ancestry and cohort groups. We found 13 Bonferroni-significant genes with an excess of PTVs and damaging missense variants, with an additional 20 genes below 5% FDR (**Figure 2b, Supplementary Table 6**). Among the most strongly associated genes were *RB1CC1* (OR = 5.82, p = 8.9 × 10^-12^), *AKAP11* (OR = 7.0, p = 2.3 × 10^-11^), and *ATP2B2* (OR = 4.1, p = 1.3 × 10^-11^). Effect sizes for Bonferroni-significant genes ranged from 2.33 to 8.68, although these estimates may be inflated by winner’s curse.

To assess potential inflation due to population structure, technical artifacts, or model misspecification, we evaluated gene-level association statistics for ultra rare synonymous variants. The synonymous CMH test showed minimal inflation (λ = 1.03), indicating appropriate calibration of the test statistics. In contrast to the analogous Fisher’s Exact Test without strata correction, the synonymous lambda was (λ = 3.01) (**Supplementary Table 6**).

Several implicated genes have been previously associated with large-scale rare variant studies in SCZ or other neurodevelopmental disorders, including *RB1CC1, HERC1, CUL1, SP4, MAGI2, AKAP11*, and *KDM5B*. Notably, multiple genes implicated here overlap those reported in the latest SCHEMA 2.0 and Autism Sequencing Consortium (ASC) studies, despite differences in sample composition and variant aggregation strategies^17,19^.

To assess enrichment across a broader range of functional annotations, we analyzed 201 BipEx genes meeting a nominal association threshold (P < 0.01) and showing increased burden in cases (OR > 1) using ToppGene^20^. This analysis identified enrichment of synaptic and postsynaptic annotations, with multiple Gene Ontology terms surviving Bonferroni correction (**Supplementary Table 7**). The strongest enrichment was observed for postsynaptic density (P_Bonf_ = 1.14 × 10^-4^; 18 of 470 annotated genes). Beyond Gene Ontology annotations, enrichment was also observed for schizophrenia-associated genes curated in DisGeNET (P_Bonf_ = 1.29 × 10^-3^; 20 of 883 genes) and insomnia-associated genes from GWAS catalogs (P_Bonf_ = 4.61 × 10^-2^; 25 of 1,620 genes), indicating overlap with genes previously implicated in psychiatric and sleep-related traits.

### Comparison of missense against PTV signal

To directly compare the relative contribution of PTVs and damaging missense variants to gene-level association signals, we plotted the gene-wise significance of missense burden against PTV burden using −log_10_(p-values) from CMH tests across ancestries (**Figure 3a, Supplementary Table 8 and 9**). A subset of genes exhibited predominantly PTV-driven signal with minimal corresponding missense association (i.e., *AKAP11*, *RB1CC1*, *SHANK1*, *KDM5B*), consistent with intolerance to loss-of-function variation. In contrast, several genes showed comparatively stronger missense signals relative to PTVs (i.e. *ATP2B2*, *EIF4E2*, *HDAC3*), suggesting sensitivity to amino acid-altering variation that may not result in complete gene disruption. Several genes showed convergence of missense and PTV, such as *SP4* and *ATP9A*; these would not have been exome-wide significant without combining both variant classes. We found that all significant genes were under strong evolutionary constraint—defined using either high sHet or low GISMO-mis scores (**Figure 3A**).

**Figure 3.**
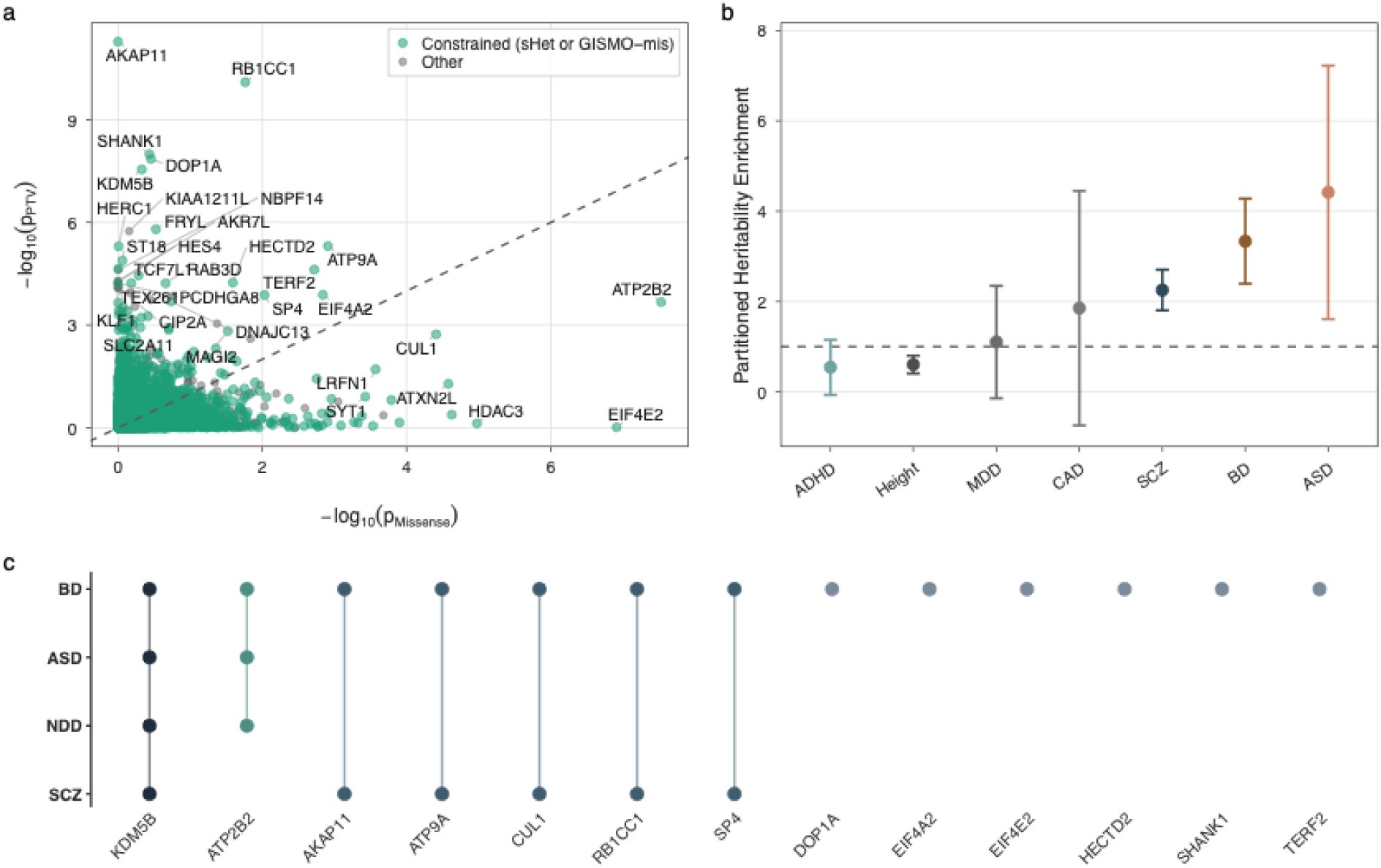
Genetic convergence across rare and common variant architectures. **(a)** Scatter plot comparing gene-level association strength for missense variants and protein-truncating variants (PTVs), shown as −log₁₀(p) values. Each point represents a gene. Genes classified as constrained based on high sHet or low GISMO-mis scores are highlighted in teal, with all other genes shown in gray. The dashed diagonal line indicates equal missense and PTV association strength. The 5% FDR significant genes are labeled. **(b)** Partitioned heritability enrichment of common variant signal within ±100 kb of the 13 Bonferroni-significant genes across multiple traits. Points denote enrichment estimates and error bars indicate ±1 standard error. The dashed horizontal line marks no enrichment (enrichment = 1). Traits are colored by category. **(c)** Overlap of Bonferroni-significant BD genes with rare-variant gene sets from SCZ, ASD, and NDD. Each column represents a BD-significant gene; filled circles indicate exome-wide significant association. Vertical connectors denote an exome-wide significant signal shared across ≥2 neurodevelopmental/psychiatric phenotypes.

### Cross-trait overlap of BipEx risk genes

To assess overlap between rare and common variant architectures, we performed partitioned heritability analyses using stratified LD score regression (S-LDSC) to test whether common variant heritability was enriched near the 13 Bonferroni-significant BD genes identified in the gene-based analysis. Gene annotations were defined as each gene body with ±100 kb flanking regions, collectively encompassing 0.14% of SNPs. We evaluated five psychiatric traits—bipolar disorder (BD), schizophrenia (SCZ), autism spectrum disorder (ASD), attention-deficit/hyperactivity disorder (ADHD), and major depressive disorder (MDD)—together with coronary artery disease (CAD) and height as non-psychiatric comparison traits. Four of five psychiatric traits showed positive effect-size estimates, with the strongest evidence of enrichment observed for SCZ (enrichment = 2.25, SE = 0.45; *P* = 0.005), which was the only trait to remain significant after Bonferroni correction across the seven traits tested (*P*Bonf = 0.037; **Figure 3b, Supplementary Table 10**). Neither non-psychiatric comparison trait showed significant positive enrichment after correction.

We additionally compared the 13 Bonferroni-significant BD genes with genes implicated by rare-variant analyses of SCZ, ASD and neurodevelopmental disorders (NDD)^17–19^. For SCZ, we included genes reaching Bonferroni-corrected significance, whereas for ASD and NDD we included genes meeting FDR < 0.001 (**Figure 3c**). Despite the small number of BD-associated genes, several overlapped genes implicated in SCZ, providing further evidence of shared genetic architecture and consistent with the literature^21,22^. Other BD-associated genes did not overlap significant SCZ, ASD, or NDD gene sets, highlighting a complementary set of associations that may reflect genetic effects more specific to BD.

### BipEx risk genes show enrichment in brain tissues, neuronal cell types, and distinct developmental expression patterns

To better understand the biological context of BipEx-associated genes, we examined their expression patterns across bulk tissues, brain cell types, and developmental stages using publicly available transcriptomic datasets. First, we assessed bulk tissue expression using GTEx v8 RNA-sequencing data^23^. Hierarchical clustering of normalized gene expression across tissues revealed that several of the 13 Bonferroni-significant BD genes showed elevated relative expression across multiple brain regions compared with non-neural tissues (**Figure 4a**). In a complementary tissue-specificity analysis, genes upregulated in the cerebellar hemisphere were significantly enriched among the 13 Bonferroni-significant BD genes (9 of 13 genes; *P* = 6.15 × 10^−4^, adjusted *P* = 0.033, **Supplementary Table 11**), and this was the only tissue-specific enrichment that remained significant after correction for multiple testing. The overlapping genes were *RB1CC1, AKAP11, ATP2B2, SHANK1, ATP9A, TERF2, SP4, HECTD2,* and *DOP1A*.

**Figure 4.**
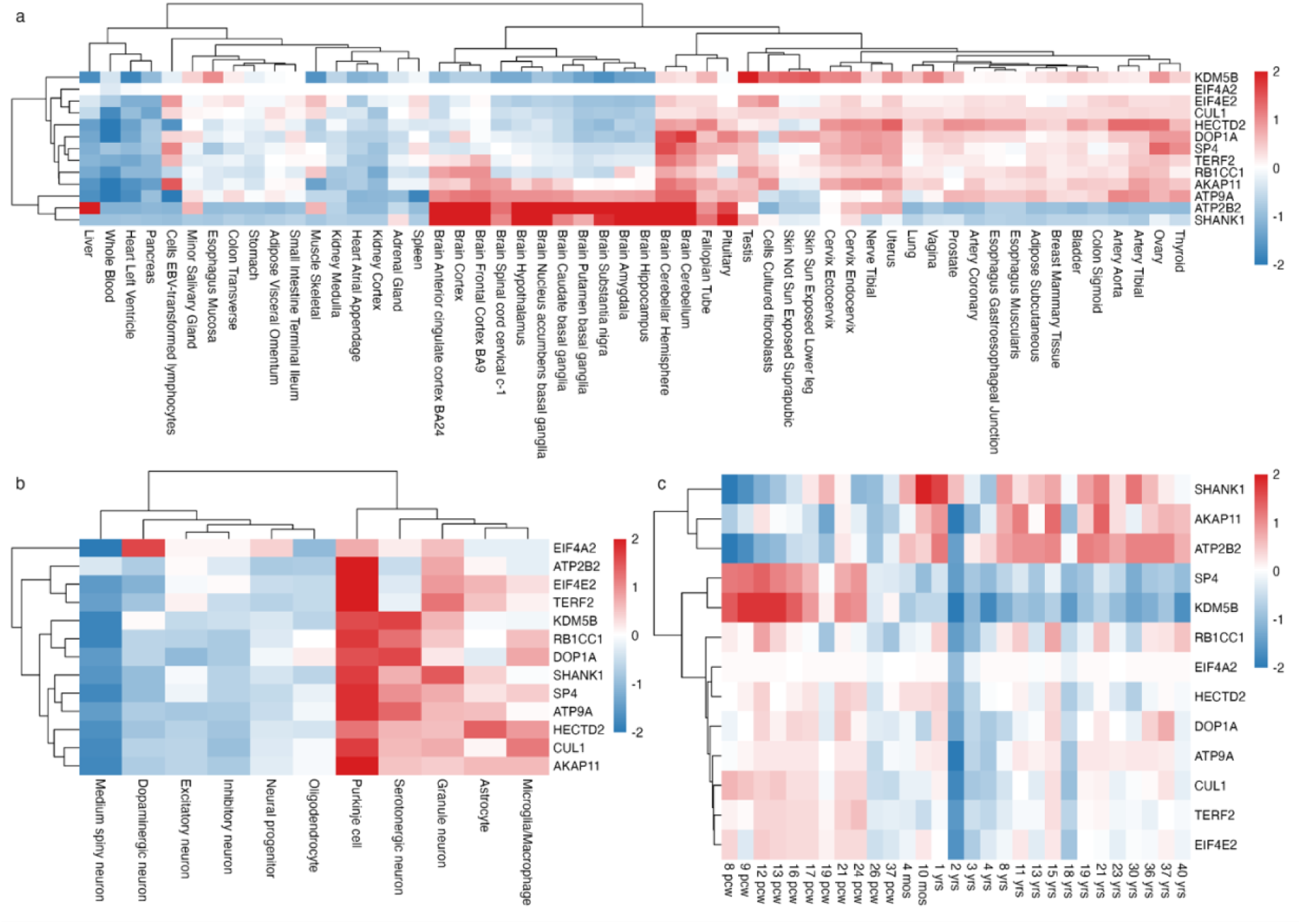
Expression profiles of BipEx risk genes across human tissues, brain cell types, and developmental stages. **(a)** Bulk tissue expression of BipEx-associated genes across human tissues from GTEx v8. Values represent normalized average expression and are scaled per gene for visualization (range truncated to –2 to 2). Both genes and tissues were hierarchically clustered using Euclidean distance. **(b)** Cell-type–resolved expression across major brain cell classes derived from CELLxGENE single-cell brain datasets. Expression values were aggregated across cell types using weighted averages and log-transformed before computing gene-wise z-scores. Rows represent genes and columns represent collapsed brain cell classes. **(c)** Developmental expression across the human lifespan from BrainSpan. Expression values correspond to average normalized RPKM across developmental stages, with genes hierarchically clustered and ages displayed in chronological order.

To investigate cell-type specificity within the brain, we next analyzed single-cell RNA sequencing data from the CELLxGENE brain atlas^24^. After collapsing individual annotations into major neuronal and glial cell classes and aggregating expression across datasets, we calculated gene-wise standardized expression profiles across cell types. Several BipEx genes showed relatively higher expression within neuronal populations compared to glial cell types (**Figure 4b**). Notably, Purkinje cells, serotonergic neurons, granule neurons, and medium spiny neurons exhibited comparatively elevated expression for multiple BipEx genes, whereas astrocytes, oligodendrocytes, and microglia generally showed lower relative expression levels (**Supplementary Table 12**). These findings suggest that BipEx-associated genes showed relatively elevated standardized expression in several neuronal populations compared with glial populations (**Supplementary Table 12**).

Finally, we examined developmental expression trajectories using BrainSpan transcriptomic data spanning prenatal development through adulthood^25^. Genes clustered into distinct developmental expression profiles (**Figure 4c**). We found that several genes, including *KDM5B* and *RB1CC1*, showed relatively higher expression during prenatal stages, whereas others demonstrated more stable expression across development or increased expression during later stages. Genes up-regulated during early childhood showed significant enrichment for BipEx genes, while other developmental windows showed weaker or non-significant enrichment signals (**Supplementary Figure 12 and 13**). These results indicate that BipEx-associated genes exhibit heterogeneous developmental expression patterns, with some genes displaying stronger prenatal expression consistent with potential roles in early neurodevelopment.

### 3D missense hotspot analysis of Bonferroni-significant BipEx genes

We next assessed whether ultra-rare missense variants exhibited spatial clustering within predicted three-dimensional protein structures of the Bonferroni-significant BipEx genes with missense signal in the CMH test. To account for potential biological and technical confounders, including ancestry and capture kit, we extended the 3D Neighborhood Test (3DNT) to incorporate covariate-adjusted case-control comparisons^26^. This extension retains the neighborhood-based enrichment framework of 3DNT while reducing the risk that localized signals reflect population structure or coverage artifacts rather than true disease enrichment (**Supplementary Figure 14**). Using this covariate-aware approach, we systematically scanned spherical regions with a 15 Å radius across each protein to identify localized enrichment of missense variants relative to the remainder of the protein.

Among the genes analyzed, *DOP1A* showed the strongest evidence of spatial missense clustering (**Figure 5a, Supplementary Table 13**). The most significant neighborhood was centered at amino acid 391 (UniProt Q5JWR5), with a raw P-value of 1.00 × 10^-10^ and was significant after multiple hypothesis testing (FDR < 0.001; FWER < 0.001) (**Supplementary Figure 15**). Interestingly, *ATP2B2*, also emerged as significantly associated, with the 15 Å radius overlapping with the calcium binding site (**Supplementary Figure 16**).

**Figure 5.**
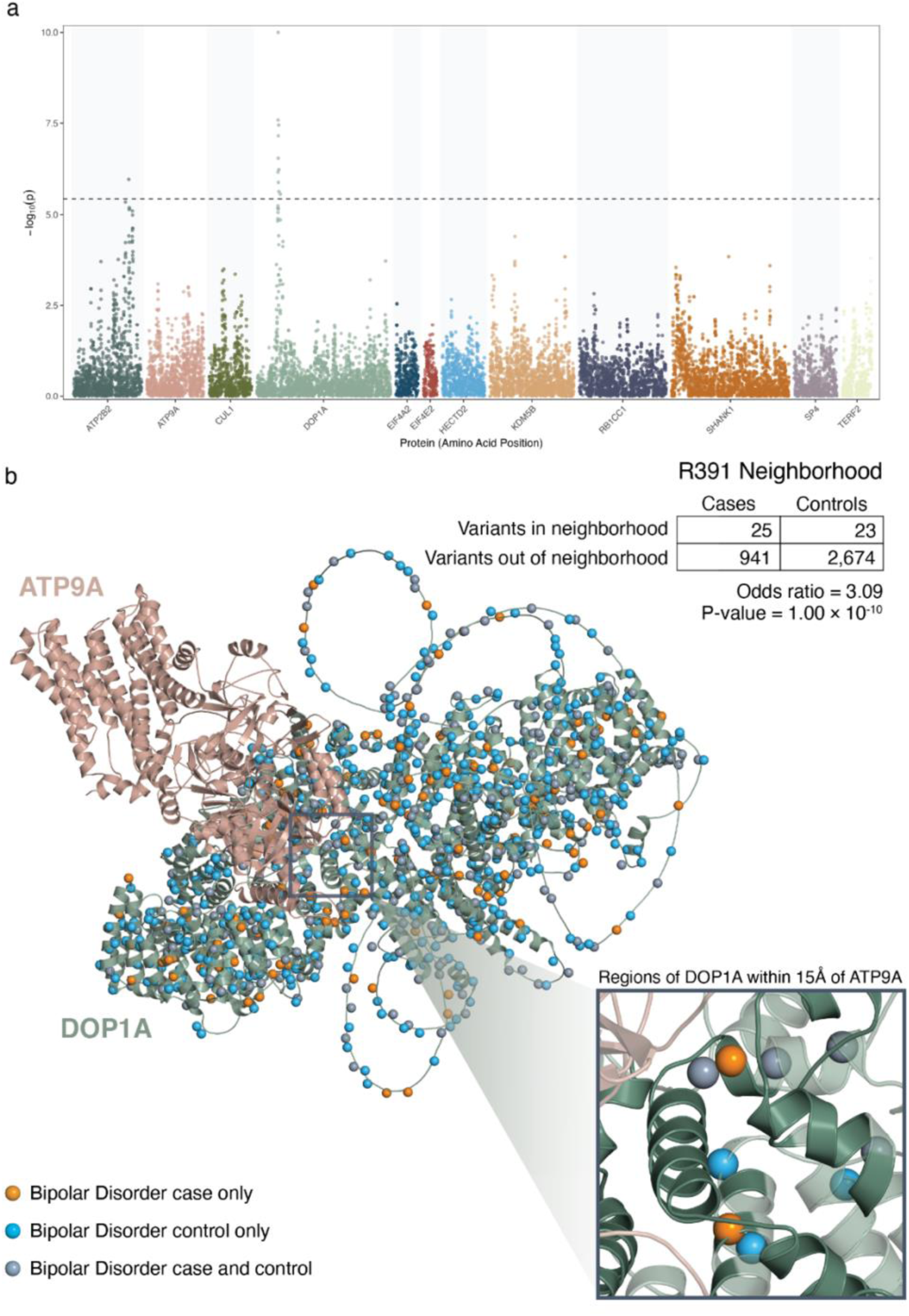
Spatial clustering of ultra-rare missense variants in the DOP1A protein structure and proximity to ATP9A interaction interface. **a**. Manhattan plot of ultra-rare 3D missense neighborhoods across Bonferroni-significant BipEx genes. Each point represents a 3D neighborhood of 15 Å radius positioned by its amino acid location within the encoded protein, with the y-axis showing the −log₁₀(P) from the covariate-adjusted 3D Neighborhood Test (3DNT). The dashed horizontal line indicates the significance threshold after correcting for multiple neighborhoods tested. DOP1A shows the strongest evidence of localized spatial clustering among the genes analyzed. The gene *AKAP11* was excluded due to no missense signal in the gene-level association testing. **b.** Structural visualization of the most significant missense clustering region in DOP1A. The 3DNT identified a neighborhood centered at residue R391 (UniProt Q5JWR5) using a 15 Å radius search, showing enrichment of missense variants in bipolar disorder cases relative to controls (odds ratio = 3.09, P = 1.00 × 10^-10^). Variants are mapped onto the predicted DOP1A protein structure, with spheres indicating case-only variants (orange), control-only variants (blue), and variants observed in both cases and controls (gray). ATP9A is shown in pink and DOP1A in green. The inset highlights the region of DOP1A located within 15 Å of ATP9A in the AlphaFold3-predicted DOP1A–ATP9A complex, demonstrating that the most strongly associated R391 hotspot lies near the predicted protein–protein interaction interface.

### Spatial clustering of missense variation identifies a candidate functional surface in *DOP1A*

To evaluate potential mechanistic relevance, we examined all protein–protein interactions predicted by STRING for *DOP1A*^27^. We assessed all interactions with the protein pairs of DOP1A in AlphaFold3 to determine whether any occurred within 15 Angstroms of the most significant neighborhood^22^, and identified a predicted interaction with ATP9A, another BipEx risk gene Structural modeling of the DOP1A–ATP9A complex using AlphaFold3 indicated that the *DOP1A* hotspot region lies near the predicted interaction surface between the two proteins (**Figure 5b**)^28^. This spatial proximity supports the possibility that clustered missense variants in *DOP1A* may perturb a specific interaction interface rather than broadly destabilizing the protein. We confirmed the AlphaFold3 prediction was of high confidence (**Supplementary Figure 17**).

Together, these results identify a discrete structural region within DOP1A, centered at residue 391, that shows significant missense enrichment and coincides with a predicted interaction surface involving ATP9A, nominating disruption of this interaction as a plausible mechanistic hypothesis.

## Discussion

The genetic architecture of severe psychiatric disorders has contributions from both widespread common and rare variation^4,6,16^. Large-scale sequencing studies have shown that ultra-rare disruptive variants can identify individual genes that substantially increase disease risk relative to common variants. Here, we demonstrate that ultra-rare protein-truncating and damaging missense variants contribute substantially to BD risk in genes under strong evolutionary constraint. Gene-level analyses identified several exome-wide significant associations, with odds ratios substantially larger (OR: 2.33–8.68) than those typically observed for common variant loci. Although these genetic variants are individually rare, the large effect sizes indicate that these ultra-rare coding variants can meaningfully increase BD liability. No significant enrichment for ultra-rare synonymous variants suggests that these associations likely are not driven by technical or population artifacts.

The results additionally point to overlap between BD and other neuropsychiatric conditions. Enrichment was observed in genes previously implicated in SCZ, ASD, and broader NDD. However, not all BD-associated genes currently show evidence of association in ASD or NDD datasets. Although these comparisons remain limited by incomplete gene discovery and differences in statistical power across disorders, the emerging pattern suggests a rare variant architecture that includes both broadly shared neuropsychiatric risk genes and potentially more disorder-enriched components. Regions surrounding the Bonferroni-significant BD genes showed significant enrichment for common-variant heritability in SCZ after correction for multiple testing. Given the small number of genes comprising the annotation, however, this result should be interpreted cautiously because S-LDSC significance testing can be poorly calibrated for small or spatially clustered annotations^29^. No significant positive enrichment was observed for the non-psychiatric comparison traits. The identification of *KDM5B* also points to a notable connection with chromatin regulation in SCZ. *SETD1A*, one of the first rare coding genes robustly implicated in SCZ^30^, encodes a component of the SET1A/COMPASS H3K4 methyltransferase complex, whereas *KDM5B* encodes an H3K4 demethylase that removes H3K4me2/3^31,32^. Thus, genetic risk for BD and SCZ presently implicates opposing regulators of the same chromatin modification. Rather than suggesting a simple directional effect of H3K4 methylation, these findings raise the possibility that disruption of H3K4 methylation homeostasis may be relevant across severe psychiatric disorders.

The direction of association was largely consistent across ancestry groups, with rare-variant enrichment in BD cases generally observed across population strata. Although some gene sets showed moderate heterogeneity in effect sizes, the overall pattern was consistent across ancestries. These results support the generalizability of the observed rare-variant associations across populations. We additionally found that pathway analyses help place these genes into a broader biological context. Genes nominally associated with BD were strongly enriched for synaptic and postsynaptic processes, including terms related to postsynaptic density, synaptic membranes, and trans-synaptic signaling. This pattern is consistent with a growing body of genetic and functional evidence linking synaptic dysfunction with psychiatric disease^33^. All in all, this suggests that disruptions in neuronal signaling and synaptic organization are potential central components of BD pathophysiology.

Furthermore, expression analyses across bulk tissues, single-cell datasets, and developmental timepoints provide further context into BD biology. The Bonferroni significant BipEx genes showed preferential expression in brain tissues and neuronal populations, with comparatively lower expression in peripheral tissues and glial cell types. Assessment of these genes in BrainSpan found significant upregulation during early childhood, a developmental period characterized by synaptic maturation and circuit refinement^34^, suggesting that BD genes may influence neuronal development and circuit formation.

Comparison of variant classes revealed substantial heterogeneity in the allelic architecture of BD risk genes. Whereas some associations were predominantly driven by protein-truncating variants, others showed stronger missense contributions or evidence from both variant classes. This heterogeneity suggests that rare coding risk cannot be explained by a single mechanism such as haploinsufficiency, and that the functional consequences of variation may differ substantially across genes. Missense-driven associations may reflect more localized mechanisms, including disruption of specific protein domains or molecular interactions, motivating gene-specific analyses of variant position and protein structure.

Among the 13 BipEx genes, we sought to assess the potential mechanisms underlying these associations. Among the genes, we found that *DOP1A* showed the strongest association for spatial clustering of ultra-rare missense variants within the predicted three-dimensional protein structure. These variants were concentrated within a compact region centered at residue 391 and suggest disruption of a localized functional site. Structural modeling with AlphaFold3 placed this region near a predicted interaction interface with *ATP9A*, and suggest the possibility that BD-associated missense variants perturb the *DOP1A*–*ATP9A* interaction and alter membrane trafficking processes. Although experimental validation will be required, these results highlight how integrating rare variant association signals with structural protein modeling can refine gene-level associations into specific mechanistic hypotheses.

There are several limitations to these analyses. Rare variant association studies rely on small numbers of carriers, and effect size estimates are therefore susceptible to winner’s curse; decreases in effect size from BipEx 1 to BipEx 2 reflect this. We caveat the interpretability of effect sizes given heterogeneity in ascertainment. Although ancestry– and platform-stratified analyses, along with extensive quality control, can reduce the likelihood of confounding, residual heterogeneity across sequencing platforms, ascertainment strategies, and diagnostic practices may still influence gene-level signals. BD is also a heterogeneous disorder with variability in severity, course and presentation, and although we worked to ensure as much diagnostic consistency across the cohort, such variation persists for studies of this scale. Composite missense pathogenicity scores also integrate predictive tools trained on datasets with ancestry imbalances, which could affect calibration across populations. In addition, pathway enrichment and 3D structural missense modelling increase biological plausibility but do not establish causality; functional validation will be necessary to determine the functional consequences of these variants.

Overall, our results support a model in which rare coding variants with large effects contribute to BD risk through constrained neuronal genes and shared biological pathways related to synaptic function and neurodevelopment. Continued growth of sequencing cohorts, together with integration of functional genomic datasets and systematic perturbation studies in human neuronal systems, will help translate these findings into biological insight. Together, these results suggest that BD risk reflects a constellation of rare and common genetic variation that converges on neuronal and synaptic biology.

## Online Methods

### Study cohorts and sequencing

BipEx 2.0 integrated coding-sequence data generated by whole-exome sequencing (WES), blended genome-exome (BGE) sequencing and whole-genome sequencing (WGS) across multiple global collections. Cohort-specific recruitment, phenotyping and ethics information are provided in the Supplementary Methods. Sequencing data included previously described BipEx 1.0 samples and additional WES, WGS and BGE datasets^6^. WES and WGS data were jointly called with gnomAD v4.0, and analyses of WGS data were restricted to exome target intervals to improve comparability across sequencing technologies. A major advance of the current study is the substantial increase in sample size relative to BipEx 1, corresponding to an approximately 6.7-fold increase in effective sample size. This expansion markedly improves power to detect rare-variant associations, including signals from genes with lower variant counts or more modest effect sizes.

BGE libraries were generated from DNA isolated from blood or saliva using the previously described BGE workflow^8^. In brief, PCR-free whole-genome libraries and exome-enriched libraries captured with Twist Alliance Clinical Research Exome probes were combined and sequenced with 2 × 150-bp reads on Illumina NovaSeq S4 (BGE-GATK) or NovaSeq X (BGE-DRAGEN) instruments. Detailed library preparation conditions are provided in the Supplementary Methods.

### Variant calling and variant-level quality control

Joint genotyping was performed using GATK or Illumina DRAGEN according to dataset provenance^35^. GATK samples were called in gVCF mode with HaplotypeCaller and jointly genotyped within callsets using GATK best-practice workflows, followed by variant quality score recalibration. DRAGEN samples were aligned and called in gVCF mode using the DRAGEN Bio-IT Platform and jointly genotyped with the DRAGEN joint-calling workflow^36^.

For WES and WGS datasets processed under the gnomAD quality-control framework, variants failing allele-specific variant quality score recalibration were excluded. Genotypes were required to have depth (DP) ≥10 and genotype quality (GQ) ≥20; heterozygous genotypes were additionally required to have allele balance (AB) ≥0.3. WGS sites were restricted to gnomAD exome target intervals. BGE-GATK and BGE-DRAGEN callsets were quality controlled separately to accommodate differences in sequencing and variant-calling workflows. Sites with more than six alternate alleles, sites outside the Twist exome target intervals, low-complexity loci and invariant sites were excluded, and multiallelic variants were decomposed before filtering. BGE-GATK variants failing AS-VQSR were removed. BGE-DRAGEN variants failing the VariantExtractTrainScore procedure were removed. For BGE-GATK, genotypes with GQ <20 or DP <10 were excluded; heterozygous calls were additionally required to have AB ≥0.3 and an informative-read fraction ≥0.8, and homozygous-alternate calls required AB ≥0.8. For BGE-DRAGEN, depth was approximated from allele-depth fields where available; heterozygous calls required GQ ≥20, estimated depth ≥10 and AB ≥0.3, while homozygous-alternate calls required GQ ≥20 and estimated depth ≥10. Additional platform-specific QC details are provided in the Supplementary Methods.

### Sample quality control, ancestry inference and relatedness

For WES and WGS data, analyses were restricted to samples passing the gnomAD sample-level quality-control framework. For BGE datasets, samples were subjected to callset-specific hard filters, ancestry assignment and outlier removal. Genetic ancestry was inferred using principal-component analysis followed by supervised random-forest classification against reference samples; gnomAD ancestry assignments were used where available. BGE ancestry inference incorporated external reference populations, and samples without a sufficiently confident ancestry assignment were excluded. Full reference-panel composition and classification procedures are provided in the Supplementary Methods.

Within cohort-by-ancestry strata, Hail sample_qc() metrics were used to identify outliers. Samples exceeding ±4 median absolute deviations (MADs) from the median for transition-to-transversion ratio, insertion and deletion counts, transition and transversion counts, total SNP count, heterozygosity ratio or indel ratio were removed. Singleton counts were filtered asymmetrically at >8 MADs above the median; an additional maximum of 5,000 singletons was applied to gnomAD v4.0 exomes. Sex concordance was evaluated using genetic-sex inference and chromosome-Y genotype counts, with discordant samples excluded. Platform-specific sex-imputation thresholds and QC distributions are shown in Supplementary Figures 1–7.

After callset-specific QC, passing samples were merged in Hail. Relatedness was estimated using KING on common, high-call-rate, LD-pruned variants within ancestry groups^37^. Independently sequenced duplicate samples were used as positive controls. Individuals with estimated relatedness >0.125 were removed to obtain a maximally independent set, preferentially retaining cases over controls when selecting among related samples. Samples with more than ten detected relationships were excluded as likely low-quality or contaminated (**Supplementary Figure 8**).

### Variant annotation and definition of qualifying variants

Variants were annotated using Ensembl Variant Effect Predictor (VEP) v95 on the Ensembl canonical transcript^10^. Transcript-ablation, splice-acceptor, splice-donor, stop-gain and frameshift variants were classified as protein-truncating variants (PTVs). High-confidence loss-of-function variants were identified using LOFTEE v1 with additional filters to remove variants in single-exon genes, variants lacking exon annotations because of intronic overlap, and terminal variants failing the 50-bp truncation rule with negative GERP^38^ distance values^39^.

Damaging missense variants were defined using an ensemble score integrating MPC v2, AlphaMissense and MisFit-S^11–13^. Each score was ranked across the exome, transformed to a Z score, and averaged to generate the MeanMis score. Variants above the 96th percentile of MeanMis were classified as damaging; this threshold was selected empirically based on case-control enrichment across the BipEx dataset (**Supplementary Figure 9**). Rare-variant analyses were restricted to variants with minor allele frequency ≤0.01%.

### Gene-based rare-variant association testing

Gene-level rare-variant association was performed using a Cochran-Mantel-Haenszel (CMH) framework stratified by sequencing platform and inferred genetic ancestry. Cohort-by-ancestry strata with fewer than 50 cases, fewer than 5 controls or fewer than 200 total samples were excluded. Association tests were conducted separately for PTVs, damaging missense variants, the combined PTV plus damaging-missense class, and synonymous variants. We selected the CMH framework after comparison with an unstratified Fisher’s exact test, which showed substantial inflation in the synonymous negative-control category, whereas the stratified CMH test remained well calibrated (**Supplementary Figure 10**).

### Gene-set burden testing

Gene-set burden analyses were performed using sample-level rare-variant burden counts aggregated across genes within each gene set and variant consequence class. PTV, damaging missense, combined PTV plus damaging-missense and synonymous burdens were tested separately. Within each major ancestry group, bipolar disorder case status was modeled by logistic regression with gene-set burden as the primary predictor and sex, the first ten ancestry principal components and sequencing platform as covariates. Analyses were restricted to ancestry strata with at least ten cases and ten controls, and tests required at least five carriers within a stratum. Ancestry-specific estimates were combined by fixed-effect inverse-variance meta-analysis on the log-odds-ratio scale. Between-ancestry heterogeneity was assessed with Cochran’s Q and I².

### Cross-disorder overlap and common-variant heritability

Cross-disorder overlap was evaluated by comparing BipEx-significant protein-coding genes with published rare-variant gene sets for schizophrenia, autism spectrum disorder and neurodevelopmental disorders^16–18^. Gene symbols were standardized to HGNC nomenclature before overlap analysis; the external significance definitions used for each disorder are detailed in the Supplementary Methods.

Partitioned heritability was estimated by stratified LD score regression (LDSC)^40^. A custom annotation was constructed from the 13 Bonferroni-significant BipEx genes, including each gene body and ±100-kb flanking regions. Stratified LDSC was applied to genome-wide association summary statistics using the baseline-LD v2.2 model as covariates and European-ancestry LD scores, with analyses restricted to HapMap3 SNPs. Enrichment was defined as the proportion of SNP heritability explained by the annotation divided by the proportion of SNPs contained in the annotation. Standard errors and P values were estimated by block jackknife, and multiple testing was controlled by Bonferroni correction. Traits analyzed included schizophrenia, bipolar disorder, autism spectrum disorder, major depressive disorder, attention-deficit/hyperactivity disorder, height and coronary artery disease^4,41–46^.

### Pathway enrichment

For pathway analyses, genes with P <0.01 in BipEx gene-level burden testing and odds ratio >1 were analyzed using ToppGene Suite (ToppFun)^20^. Enrichment was evaluated across Gene Ontology biological-process and cellular-component categories, curated disease annotations including DisGeNET and the GWAS Catalog, and PubMed-derived gene sets. Hypergeometric tests were used to compare the input gene list with the genomic background. Multiple-testing correction included Bonferroni and false-discovery-rate procedures; pathways highlighted in the main text were required to pass Bonferroni correction.

### Transcriptomic analyses

Expression analyses focused on the 13 BipEx-associated genes *ATP2B2, SP4, RB1CC1, SHANK1, DOP1A, KDM5B, ATP9A, CUL1, EIF4E2, EIF4A2, HECTD2, TERF2 and AKAP11*.

Bulk-tissue and developmental expression were evaluated using GTEx v8 and BrainSpan datasets through FUMA gene2func^23,25,47^. Gene-by-tissue or gene-by-age expression matrices were generated using standardized HGNC symbols, and hierarchical clustering used Euclidean distance with complete linkage. FUMA differential-expression enrichment analyses were used to test whether BipEx genes were overrepresented among genes up– or downregulated in specific tissues, ages or developmental stages (**Supplementary Figures 11–13**).

Single-cell expression was evaluated using CELLxGENE brain gene-by-cell-type summaries^24^. Analyses were restricted to brain tissue, and non-brain or clearly irrelevant annotations were excluded. Fine-grained annotations were collapsed by rule-based mapping into canonical neuronal, glial and vascular cell classes. For each gene and collapsed class, total cell counts, numbers of expressing cells, cell-count-weighted mean expression and percentage of expressing cells were calculated; classes with fewer than 200 cells were excluded. Absolute expression was visualized by dot plots using percent expressing and log1p-transformed mean expression, and relative expression was summarized using within-gene Z scores. Detailed annotation rules, missingness filters and visualization procedures are provided in the Supplementary Methods.

### Three-dimensional missense neighborhood analysis

Spatial clustering of disease-associated missense variants was evaluated using the 3D Neighborhood Test (3DNT) in the 13 BipEx genes reaching exome-wide significance^26^. Ultra-rare missense variants were mapped to canonical protein sequences and corresponding three-dimensional structures using precomputed genomic-to-protein mappings and AlphaFold-predicted structures; AlphaFold3 models were generated when structures were unavailable in the AlphaFold Protein Structure Database^28^. Variants that could not be confidently mapped were excluded.

For each protein residue, a spherical neighborhood with a 15-Å radius was defined, excluding residues with predicted aligned error >15 Å relative to the neighborhood center. Association between neighborhood membership and case-control status was tested by logistic regression. The model included a sample-specific covariate representing the baseline probability of case status derived from case-control counts within the corresponding cohort-by-ancestry stratum. To account for the large number of correlated neighborhood tests, empirical false-discovery rates were estimated from 1,000 null simulations and constrained to be monotonic across P-value thresholds. Neighborhoods with empirical FDR <0.05 were considered significant. Comparison with a Fisher’s exact framework and structural association maps are shown in **Supplementary Figures 14–16**.

### Protein-interaction and structural modeling

To contextualize significant missense neighborhoods, first-degree protein interactors for each BipEx gene were retrieved from STRING v11.5 using default confidence thresholds, retaining interactions supported by experimental evidence, curated databases or strong coexpression^27^. STRING networks were used descriptively rather than for formal enrichment testing.

Selected high-confidence interacting protein pairs were modeled with AlphaFold3 to evaluate whether significant 3DNT neighborhoods overlapped predicted protein-protein interfaces. Predicted interfaces were assessed on the basis of cross-protein residue proximity together with AlphaFold3 confidence metrics. Structural overlap was used to assess whether clustered disease-associated missense variation localized to predicted interaction surfaces (**Supplementary Figure 17**).

## Data Availability

All data produced in the present work are contained in the manuscript and available on https://bipex.broadinstitute.org/.

## Acknowledgements

C.L. is supported by the NIMH K99MH141141. M.A. Stanley Center, NIMH R01MH120642, NIMH U01MH125047. S.G. Stanley Center, NIMH U01MH125047. R.M.M. Stanley Center. C.R.J.C.N.

Stanley Center. Z.Z. Stanley Center. M.L. We are deeply grateful for the participation of all subjects contributing to this research, and to the collection team that worked to recruit them, including the Swedish National Quality Register for bipolar disorder (BipoläR). Funding support for SWEBIC was provided by the Stanley Center for Psychiatric Research, Broad Institute from a grant from Stanley Medical Research Institute (Subaward No. 5710002350), the Swedish Research Council (2022-01643), the Swedish Foundation for Strategic Research (KF10-0039), and the Swedish Brain Foundation (FO2025-0004-HK-212). P.-H.K. This study was funded by National Science and Technology Council (NSTC 108-2314-B-002-136-MY3, 110-2314-B-002-067-MY3) and Population Health Research Center from Featured Areas Research Center Program within the framework of the Higher Education Sprout Project by the Ministry of Education in Taiwan (grant number NTU-112L9004). S.E.M. The Australian Genetics of Bipolar Disorder Study (GBP) data collection was funded and data analysis was supported by an Australian National Health and Medical Research Council Grant (No. APP1138514) to S.E.M. S.E.M. is supported by a NHMRC Investigator Grant (No. APP2025674). We thank the participants for giving their time and support for this project. We acknowledge and thank M. Steffens for her generous donations and fundraising support. L.A. Stanley Center, NIMH U01MH125047. A.D. Stanley Center, NIMH R01MH120642, NIMH U01MH125047. S.M.K. Stanley Center. S.T. Stanley Center, NIMH R01MH120642, NIMH U01MH125047. D.J.S. Stanley Center, NIMH R01MH120642. H.H. H.H. acknowledges support from Merkin Institute Fellow and the National Institute of Diabetes and Digestive and Kidney Diseases (nos K01DK114379 and R01DK129364). H. Huang. Was funded by the Merkin Institute Fellow, the Zhengxu and Ying He Foundation, Stanley Center, NIMH R01MH130675. This research was funded in part by BD2: Breakthrough Discoveries for Thriving with Bipolar Disorder. For the purpose of open access, the author has applied a CC BY 4.0 public copyright license to all Author Accepted Manuscripts arising from this submission. B.M.N. This research was funded in part by BD2: Breakthrough Discoveries for Thriving with Bipolar Disorder. For the purpose of open access, the author has applied a CC BY 4.0 public copyright license to all Author Accepted Manuscripts arising from this submission. This research was also supported by R01MH101244, R37MH107649, U01MH125047, and U24MH140953. The authors note with sadness that D.J.S. (Dan J. Stein) passed away during the preparation of this manuscript.

## References

1. Carvalho, A. F., Firth, J. & Vieta, E. Bipolar Disorder. N. Engl. J. Med. 383, 58–66 (2020).

2. Family, twin, and adoption studies of bipolar disorder – Smoller – 2003 – American Journal of Medical Genetics Part C: Seminars in Medical Genetics – Wiley Online Library. https://onlinelibrary.wiley.com/doi/10.1002/ajmg.c.20013.

3. Kendler, K. S., Ohlsson, H., Sundquist, J. & Sundquist, K. An Extended Swedish National Adoption Study of Bipolar Disorder Illness and Cross-Generational Familial Association With Schizophrenia and Major Depression. JAMA Psychiatry 77, 814–822 (2020).

4. O’Connell, K. S. et al. Genomics yields biological and phenotypic insights into bipolar disorder. Nature 639, 968–975 (2025).

5. Mullins, N. et al. Genome-wide association study of more than 40,000 bipolar disorder cases provides new insights into the underlying biology. Nat. Genet. 53, 817–829 (2021).

6. Palmer, D. S. et al. Exome sequencing in bipolar disorder identifies AKAP11 as a risk gene shared with schizophrenia. Nat. Genet. 54, 541–547 (2022).

7. Thorgeirsson, T. E. et al. Rare loss-of-function variants in HECTD2 and AKAP11 confer risk of bipolar disorder. Nat. Genet. 57, 851–855 (2025).

8. Boltz, T. A. et al. A blended genome and exome sequencing method captures genetic variation in an unbiased and cost-effective manner. Nat. Genet. 1–9 (2026) doi:10.1038/s41588-026-02669-w.

9. Martin, A. R. et al. Clinical use of current polygenic risk scores may exacerbate health disparities. Nat. Genet. 51, 584–591 (2019).

10. McLaren, W. et al. The Ensembl Variant Effect Predictor. Genome Biol. 17, 122 (2016).

11. Accurate proteome-wide missense variant effect prediction with AlphaMissense | Science. https://www.science.org/doi/10.1126/science.adg7492.

12. Wang, L. et al. The landscape of regional missense mutational intolerance quantified from 730,947 exomes. 2024.04.11.588920 Preprint at 10.1101/2024.04.11.588920 (2026).

13. Zhao, Y. et al. A probabilistic graphical model for estimating selection coefficients of nonsynonymous variants from human population sequence data. Nat. Commun. 16, 4670 (2025).

14. Liao, C. et al. The landscape of gene loss and missense variation across the mammalian tree informs on gene essentiality. 2024.05.16.594531 Preprint at 10.1101/2024.05.16.594531 (2024).

15. Zeng, T., Spence, J. P., Mostafavi, H. & Pritchard, J. K. Bayesian estimation of gene constraint from an evolutionary model with gene features. Res. Sq. rs.3.rs-3012879 (2023) doi:10.21203/rs.3.rs-3012879/v1.

16. Singh, T. et al. Rare coding variants in ten genes confer substantial risk for schizophrenia. Nature 604, 509–516 (2022).

17. Fu, J. M. et al. Rare coding variation provides insight into the genetic architecture and phenotypic context of autism. Nat. Genet. 54, 1320–1331 (2022).

18. Kaplanis, J. et al. Evidence for 28 genetic disorders discovered by combining healthcare and research data. Nature 586, 757–762 (2020).

19. Schema, C. Analysis of Rare Coding Variation Identifies New Genetic Contributors to Schizophrenia.

20. Chen, J., Bardes, E. E., Aronow, B. J. & Jegga, A. G. ToppGene Suite for gene list enrichment analysis and candidate gene prioritization. Nucleic Acids Res. 37, W305–W311 (2009).

21. Grotzinger, A. D. et al. Mapping the genetic landscape across 14 psychiatric disorders. Nature 649, 406–415 (2026).

22. Cross-Disorder Group of the Psychiatric Genomics Consortium et al. Genetic relationship between five psychiatric disorders estimated from genome-wide SNPs. Nat. Genet. 45, 984–994 (2013).

23. GTEx Consortium. The Genotype-Tissue Expression (GTEx) project. Nat. Genet. 45, 580– 585 (2013).

24. Abdulla, S. et al. CZ CELLxGENE Discover: a single-cell data platform for scalable exploration, analysis and modeling of aggregated data. Nucleic Acids Res. 53, D886–D900 (2024).

25. Kang, H. J. et al. Spatio-temporal transcriptome of the human brain. Nature 478, 483–489 (2011).

26. Nason, E. et al. Systematic identification of disease-associated 3D neighborhoods in protein structures. 2026.05.29.26354366 Preprint at 10.64898/2026.05.29.26354366 (2026).

27. Szklarczyk, D. et al. The STRING database in 2023: protein–protein association networks and functional enrichment analyses for any sequenced genome of interest. Nucleic Acids Res. 51, D638–D646 (2023).

28. Abramson, J. et al. Accurate structure prediction of biomolecular interactions with AlphaFold 3. Nature 630, 493–500 (2024).

29. Tashman, K. C., Cui, R., O’Connor, L. J., Neale, B. M. & Finucane, H. K. Significance testing for small annotations in stratified LD-Score regression. 2021.03.13.21249938 Preprint at 10.1101/2021.03.13.21249938 (2021).

30. Singh, T. et al. Rare loss-of-function variants in SETD1A are associated with schizophrenia and developmental disorders. Nat. Neurosci. 19, 571–577 (2016).

31. Xiang, Y. et al. JARID1B is a histone H3 lysine 4 demethylase up-regulated in prostate cancer. Proc. Natl. Acad. Sci. U. S. A. 104, 19226–19231 (2007).

32. Kristensen, L. H. et al. Studies of H3K4me3 demethylation by KDM5B/Jarid1B/PLU1 reveals strong substrate recognition in vitro and identifies 2,4-pyridine-dicarboxylic acid as an in vitro and in cell inhibitor. FEBS J. 279, 1905–1914 (2012).

33. Göteson, A. et al. Cerebrospinal fluid biomarkers reveal transdiagnostic synaptic dysfunction across major psychiatric disorders. Nat. Commun. 17, 7604 (2026).

34. Huttenlocher, P. R. & Dabholkar, A. S. Regional differences in synaptogenesis in human cerebral cortex. J. Comp. Neurol. 387, 167–178 (1997).

35. McKenna, A. et al. The Genome Analysis Toolkit: A MapReduce framework for analyzing next-generation DNA sequencing data. Genome Res. 20, 1297–1303 (2010).

36. Behera, S. et al. Comprehensive genome analysis and variant detection at scale using DRAGEN. Nat. Biotechnol. 43, 1177–1191 (2025).

37. Manichaikul, A. et al. Robust relationship inference in genome-wide association studies. Bioinformatics 26, 2867–2873 (2010).

38. Davydov, E. V. et al. Identifying a high fraction of the human genome to be under selective constraint using GERP++. PLoS Comput. Biol. 6, e1001025 (2010).

39. Karczewski, K. J. et al. The mutational constraint spectrum quantified from variation in 141,456 humans. Nature 581, 434–443 (2020).

40. Bulik-Sullivan, B. K. et al. LD Score regression distinguishes confounding from polygenicity in genome-wide association studies. Nat. Genet. 47, 291–295 (2015).

41. Trubetskoy, V. et al. Mapping genomic loci implicates genes and synaptic biology in schizophrenia. Nature 604, 502–508 (2022).

42. Grove, J. et al. Identification of common genetic risk variants for autism spectrum disorder. Nat. Genet. 51, 431–444 (2019).

43. Major Depressive Disorder Working Group of the Psychiatric Genomics Consortium. Electronic address: & Major Depressive Disorder Working Group of the Psychiatric Genomics Consortium. Trans-ancestry genome-wide study of depression identifies 697 associations implicating cell types and pharmacotherapies. Cell 188, 640–652.e9 (2025).

44. Demontis, D. et al. Genome-wide analyses of ADHD identify 27 risk loci, refine the genetic architecture and implicate several cognitive domains. Nat. Genet. 55, 198–208 (2023).

45. Yengo, L. et al. A saturated map of common genetic variants associated with human height. Nature 610, 704–712 (2022).

46. Aragam, K. G. et al. Discovery and systematic characterization of risk variants and genes for coronary artery disease in over a million participants. Nat. Genet. 54, 1803–1815 (2022).

47. Watanabe, K., Taskesen, E., van Bochoven, A. & Posthuma, D. Functional mapping and annotation of genetic associations with FUMA. Nat. Commun. 8, 1826 (2017).

