## Supplementary Text and Figures for "Rare coding variation implicates thirteen genes in bipolar disorder across 232,536 individuals from global populations"

### Supplementary Methods and Figures

#### Cohort descriptions

**Misión Origen**

Misión Origen is an EHR-linked biobank comprising more than 93,000 individuals with serious mental illness and population controls from the Paisa region of Colombia. Developed through a long-standing collaboration between UCLA and Universidad de Antioquia, Misión Origen builds on earlier research in the Paisa population to enable large-scale investigation of the genetic architecture and longitudinal clinical trajectories of serious mental illness. Cases were recruited through four psychiatric hospitals that provide care to a large proportion of the region, while controls were recruited through primary care clinics affiliated with SURA, one of Colombia's largest health insurance and health care providers. Available electronic health records (EHRs) span up to two decades and include demographic information, diagnoses and other clinical codes, medications, laboratory results, clinical encounters, and unstructured clinical notes. Misión Origen also incorporates the Paisa Cohort, a deeply phenotyped sample of 9,107 participants with extensive clinical, cognitive, and transdiagnostic symptom data.

**Paisa Cohort**

The Paisa Cohort is a cohort of 9,107 adults that was established prior to, and subsequently incorporated into, the broader Misión Origen biobank. The cohort was designed to investigate the genetic basis of serious mental illness in the Paisa population of Colombia, a genetic isolate of approximately 9 million individuals descended from a relatively small founder population and shaped by rapid population expansion following a demographic bottleneck during the 16th and 17th centuries (Service et al. 2006; Bedoya et al. 2006; Mooney et al. 2018). The cohort includes individuals with schizophrenia, schizoaffective disorder, bipolar disorder, or severe recurrent major depressive disorder, as well as screened community controls. Participants underwent a deep phenotyping protocol, including extensive clinical, cognitive, and transdiagnostic symptoms assessment, complemented by longitudinal EHR data. The cohort design, recruitment procedures, and phenotypic assessments have been described previously (Service et al. 2020; Lopera-Maya et al. 2026; Ramirez-Diaz et al. 2024).

**PUMAS Colombia**

The *"Populations Underrepresented in Mental Illness Association Studies (PUMAS)"* project was established with the overarching goal of building a large sample bank of serious mental illness cases and controls from Africa and admixed populations in the Americas, to ultimately increase the representation of non-European populations in psychiatric genetics research. Through a collaboration between UCLA and Universidad de Antioquia, PUMAS Colombia recruited 5,105 participants from two geographically and ancestrally distinct areas: the departments of Antioquia and Caldas in the Andean region, and Cartagena on the Caribbean coast.

In Antioquia and Caldas, the cohort included participants with serious mental illness and population controls recruited from non-Paisa populations, with phenotyping based on longitudinal EHR. In Cartagena, cases were recruited through the CEMIC psychiatric hospital and characterized through direct clinical assessments; no controls were recruited at this location. Recruitment in Cartagena was designed to capture the distinct three-way admixed ancestry of Colombia's Caribbean coastal population, with African, Indigenous American, and European ancestry components.

**PUMAS Brazil**

PUMAS Brazil was developed through a collaboration among UCLA, Universidad de Antioquia, and the Universidade Federal de São Paulo. The cohort includes over 14,000 participants with mood or psychotic spectrum disorders and screened controls recruited across five major Brazilian cities: São Paulo, Fortaleza, Salvador, Goiânia, and Belém. These cities represent populations that differ in their cultural, geographic, and ancestral backgrounds. Recruitment methods and cohort characteristics have been described previously (Carneiro et al. 2026; Ramirez-Diaz et al. 2024)

**PUMAS Los Angeles**

PUMAS Los Angeles includes 1,625 Hispanic or Latino participants with mood or psychotic spectrum disorders recruited across Los Angeles County. The cohort uses EHR-based recruitment and phenotyping at Harbor-UCLA Medical Center, Olive View-UCLA Medical Center, and San Fernando Mental Health Center. Available longitudinal data include demographic information, diagnoses and other clinical codes, medications, laboratory results, health care encounters, and unstructured clinical notes.

#### Taiwan

As part of the Stanley Center's mission to expand our global collections, we collaborated with Professor Po-Hsiu Kuo and team from the National Taiwan University (NTU) to bring in DNA samples from this Taiwanese cohort and generate sequence data through the Blended Genome Exome pipeline.

#### Whole Genome Sequencing of Psychiatric Disorders (WGSPD)

The WGSPD study sought to increase the discovery of common and low frequency variants and rare coding variants found to be associated with the risk of bipolar disorder (BD) and schizophrenia (SZ) beyond individuals of European genetic ancestry, which had primarily been the focus of previous efforts. The group performed whole genome sequencing of African American and African ancestry individuals. Data from the WGSPD grant were incorporated into the BipEx meta-analysis.

#### A-BIG-NET

A-BIG-NET is a large multi-site research study, planning to collect over 30,000 bipolar cases and controls in five Asia countries - Taiwan, Singapore, Korea, India, and

Vietnam. The study will collect a rich array of phenotyping information in Asian population.

**BioX**
The BioX project is a collaboration between Stanley Center and Shanghai Jiao Tong University in China. Through the large-scale data analysis, we aim to expand knowledge of the genetic basis of schizophrenia in Asian populations, identify genetic factors driving schizophrenia outside of European ancestries, compare schizophrenia genetic risk factors across major world populations, and ensure that the advances in genetic understanding are not simply restricted to Europeans. The controls were used in this study from BioX.

**Genomic Psychiatry Cohort (GPC)**
The Genomic Psychiatry Cohort (GPC) is a multi-institutional collaboration led by Rutgers University. The GPC resource includes an NIMH-managed repository of genomic samples at SAMPLED, genotypic and sequence data, and detailed clinical and demographic data for investigations of schizophrenia, bipolar disorder, OCD, and COVID from a variety of ancestries collected in the USA.

**GenBLIP**
The GenBLIP program recruits in Pakistan to expand the global collections and focus on discovery of additional genetic variants associated with bipolar disorder with the goal of collecting 12,000 controls and 10,000 BD cases. We have formed a consortium of Pakistani psychiatrists at 11 centers. There is a strong tradition of consanguineous marriages in Pakistan which has an advantage for genetic studies, especially of recessively inherited traits.

**Neuropsychiatric Genetics in African Populations (NeuroGAP-Psychosis)**
The Stanley Center at the Broad Institute of MIT & Harvard initiated the NeuroGAP-Psychosis project in 2015, a collaboration with colleagues at Addis Ababa University in Ethiopia, KEMRI-Wellcome Trust in Kenya, Makerere University in Uganda, Moi University/Moi Teaching and Referral Hospital in Kenya, the University of Cape Town in South Africa, and the Harvard T.H. Chan School of Public Health in the United States. With initial plans to recruit 35,000 participants (half cases with a diagnosis of schizophrenia or bipolar disorder and half controls), the target was expanded to 39,000 participants via the “Populations Underrepresented in Mental Illness Association Studies” (PUMAS) project awarded by NIMH. Sample recruitment ultimately exceeded the revised target, with >42,000 samples collected across the five NeuroGAP-Psychosis collection sites.

**NeuroMex**

The NeuroMex study is a collaboration between the Broad Institute, Harvard T.H. Chan School of Public Health, and the Ramón de la Fuente Muñiz National Institute of Psychiatry (INPRFM) in Mexico. The study aims to expand knowledge of the genetic and environmental risk factors for neuropsychiatric disorders in Mexico through large-scale sample collection and analysis, so that future advances in science and therapeutics can account for and be applicable to Mexican populations.

#### QIMR

The QIMR cohort comprises cases from the Australian Genetics of Bipolar Disorder Study (GBP) and controls from the QSkin Sun and Health Study (QSkin). The GBP study is a nation-wide cohort of 6,682 adults who self-reported experiencing bipolar disorder in their lifetime. The study aims to discover the specific and heritable genes associated with risk of bipolar disorder and response to treatment. In 2011, the QSkin study recruited 43,794 Queenslanders aged 40–69 years randomly sampled from the population of Queensland, Australia to investigate the development of skin cancer and melanoma. Self-reported diagnoses of mental health disorders were collected, including bipolar disorder, schizophrenia, and depression.

**Ethics approval** for all aspects of the project for GBP (P3408) and QSkin (P1309) was obtained from the QIMR Berghofer Human Research Ethics Committee.

**Funding**: The GBP study acknowledges and thanks M. Steffens for her generous donations in loving memory of J. Banks. Data collection was funded and data analysis was supported by the National Health and Medical Research Council (NHMRC) of Australia (No. APP1138514) to S.E.M.. D.J.S. is supported by a NHMRC Investigator Grant (No. APP1194635). S.E.M. is supported by a NHMRC Investigator Grant (No. APP2025674). The QSkin study was supported by grants from the NHMRC (APP1073898; APP1063061; APP1185416). D.C.W. is supported by an NHMRC Investigator Grant (APP2026567).

**Acknowledgements**: The GBP study thanks the participants for giving their time and support for this project. We thank David Whiteman and Catherine Olsen for their ongoing leadership of the QSkin Sun and Health Study and contributing controls to these analyses. We would also like to thank QSkin participants for giving their time and support for this project.

### Data production

#### Sequencing and alignment

In BipEx 2.0, we sought to integrate exome data from 3 separate technologies: whole-exome sequencing (WES), blended-genome exome (BGE) sequencing and whole-genome sequencing. Both WES and genomes were jointly called externally with gnomAD v4.0. All data were sequenced at the Broad Institute. Sequencing data from BipEx 1.0 were described previously in Palmer et al. (2022).

#### Blended Genome-Exome Library Preparation and Sequencing

DNA utilized for the BGE workflows (BGE-GATK and BGE-DRAGEN) was isolated from saliva and peripheral blood samples by external laboratories. Post-extraction processing followed the BGE methodology described previously (Boltz et al., 2025). In brief, genomic DNA concentrations were first adjusted to 50 ng/μL and aliquoted into 384-well plates, then purified using a 2.75× SPRI bead cleanup with AMPure XP. Concentrations were measured via spectrophotometry and samples were subsequently diluted to 25 ng/μL. A total input of 134 ng per sample was used for PCR-free library construction with customized NEBNext Ultra II FS reagents, applying reduced fragmentation, end-repair, and A-tailing conditions (37 °C for 42.57 min followed by 65 °C for 30 min).

Libraries were ligated to unique dual-index adapters at 20 °C for 20 min and underwent dual SPRI size selection steps (0.5× then 0.55×). Quantification was performed using qPCR, after which libraries were normalized to approximately 0.3–2 nM, pooled, and concentrated.

For exome sequencing, pooled PCR-free libraries were aliquoted and amplified by PCR (12 cycles; 98 °C denaturation and 65 °C annealing/extension), followed by purification with a 1× SPRI cleanup. Amplified libraries were normalized to 70 ng/μL and combined for hybrid capture using Twist Alliance Clinical Research Exome probes according to the xGen capture workflow.

PCR-free whole-genome and exome-enriched pools were quantified within the same qPCR run, mixed at a 67:33 WGS:WES ratio, and quantified again prior to sequencing. Final pools of uniquely indexed libraries were sequenced using 2×150 bp reads on either an Illumina NovaSeq S4 platform (BGE-GATK) or an Illumina NovaSeq X system (BGE-DRAGEN).

#### Variant joint calling

Joint genotyping for BipEx samples was performed using either the Genome Analysis Toolkit (GATK) or Illumina DRAGEN workflows, depending on dataset provenance (BGE-GATK vs. BGE-DRAGEN).

For samples processed with GATK, individual BAM files first underwent local realignment around indels and base quality score recalibration. Variant discovery was then conducted per sample using HaplotypeCaller in gVCF mode, generating genome-wide records containing genotype likelihoods for both variant and reference positions. Within each GATK-derived dataset (gnomAD exomes, gnomAD genomes, Cardiff/UCLA, and BGE-GATK), sample-level files were merged and jointly genotyped according to GATK best practices, using CombineVCFs for aggregation and

GenotypeVCFs for joint calling with default parameters. Variant quality annotation and filtering were subsequently applied using Variant Quality Score Recalibration (VQSR).

For samples processed with Illumina DRAGEN, sequence alignment and variant detection were carried out using the DRAGEN Bio-IT Platform, which integrates hardware-accelerated modules for read mapping, duplicate marking, indel realignment, and base quality recalibration within a unified pipeline. Variant calling was performed on each sample in gVCF mode to retain likelihood information across both variant and invariant genomic sites. For the DRAGEN-derived dataset (BGE-DRAGEN), per-sample gVCFs were combined and jointly genotyped using the DRAGEN joint-calling workflow under recommended default parameters.

#### Variant-level quality control

##### Whole exome and genome sequencing

Exome and whole-genome sequencing cohorts were obtained from gnomAD release v4.0. All selected samples were processed under the gnomAD quality control framework as described in Guez et al. (2026). To ensure comparability between platforms, whole-genome variant sites were restricted to the gnomAD exome target intervals.

Following cohort subsetting, allele-specific Variant Quality Score Recalibration (AS-VQSR) was applied independently to exome and genome datasets, and variants that did not pass recalibration thresholds were excluded. Genotype-level filtering retained only adjusted calls meeting the following criteria: sequencing depth (DP) ≥ 10, genotype quality (GQ) ≥ 20, and heterozygous allele balance (AB) ≥ 0.3. We opted for a stricter AB than 0.20 given the focus on ultra-rare variants and the possibility of introducing somatic mutations at lower AB.

##### Blended genome-exome (GATK) data

For the NeuroGAP and partially for NeuroMex datasets, raw sequencing data were accessed rather than pre-filtered releases. Comprehensive quality control was therefore performed to define high-confidence variant sites, genotypes, and samples. Because sequencing workflows and joint-calling procedures differed across datasets, quality control was conducted independently for each callset.

Variant- and genotype-level filters were applied as follows. At the variant level, sites were excluded if they contained more than six alternate alleles, fell outside Twist exome capture target intervals with 50 bp padding, or overlapped low-complexity regions. We added an additional 50 bp padding to ensure no essential splice variants get unintentionally dropped. Multiallelic loci were decomposed into biallelic records prior to filtering. Variants failing allele-specific Variant Quality Score Recalibration (AS-VQSR) were removed, as were invariant sites.

Genotype-level filtering was harmonized across callsets. Genotypes were removed if genotype quality (GQ) was <20 or sequencing depth (DP) was <10. Among heterozygous calls, genotypes were excluded if allele balance (AB) was < 0.3 or if the proportion of informative reads—defined as (reference allele depth + alternate allele depth) divided by total depth—was < 0.8. Homozygous alternate calls were excluded when AB was < 0.8. We opted for a stricter AB than 0.20 given the focus on ultra-rare variants and the possibility of introducing somatic mutations at lower AB.

##### Blended genome-exome (DRAGEN) data

A subsequent version of the Blended Genome–Exome product was processed using Illumina's DRAGEN platform (BGE-DRAGEN). The DRAGEN pipeline implements multigenome alignment against pangenome reference panels and machine learning–based variant calling within a computationally efficient framework. DRAGEN outputs are distributed via the Genomic Variant Store (GVS) rather than the Hail Variant Dataset (VDS). Because GVS omits explicit read-depth fields and caps genotype quality values at 50, several quality control procedures used for BGE-GATK were not directly transferable.

GVS data were first converted into a Hail MatrixTable format for downstream processing. Variant-level filtering excluded sites with more than six alternate alleles, positions outside Twist exome capture target intervals, and loci overlapping low-complexity regions. Multiallelic variants were decomposed into biallelic records prior to filtering. Variants failing the VariantExtractTrainScore (VETS) procedure—DRAGEN's functional analogue to GATK Variant Quality Score Recalibration—were removed, as well as invariant loci.

To approximate sequencing depth, allele depths from the allele balance field were summed for heterozygous and homozygous alternate genotypes. Homozygous reference calls in DRAGEN outputs do not provide allele balance or depth metrics; therefore, genotype-level quality control required homozygous reference genotypes to have genotype quality (GQ) ≥ 20. For heterozygous genotypes, calls were excluded if GQ < 20, estimated depth < 10, or allele balance < 0.3. Homozygous alternate genotypes were removed if GQ < 20 or estimated depth < 10.

#### Population inference

Following variant level quality control, we merged all passing samples from the different cohorts into a single Hail MatrixTable. Sample population inferences for gnomAD samples were taken from the gnomAD quality control framework. To infer sample population among BGE samples, we focused on variants with a callrate >0.99 and a minor allele frequency greater than 1%. On the combined dataset, we filtered to LD-pruned variants part of the gnomAD v4.0 PCA loadings. A random forest classifier was trained on the gnomAD v4.0 reference panel samples using continental ancestry labels and the top principal components as predictors. The trained model was applied to BGE samples, and ancestry labels were assigned only when posterior probability for a single ancestry exceeded 70%. Samples not meeting this confidence threshold were excluded. A random forest classifier was trained on reference panel samples using continental ancestry labels and the top principal components as predictors. The trained model was applied to BGE samples, and ancestry labels were assigned only when posterior probability for a single ancestry exceeded 70%. Samples not meeting this confidence threshold were excluded from analysis.

#### Sample-level quality control

##### Whole exome and whole genome sequencing

For the exome and genome data, we restricted analyses to samples designated as high quality following completion of the gnomAD sample-level quality control pipeline. This pipeline excluded individuals failing predefined hard filters, including excessive contamination, abnormal singleton counts, extreme heterozygosity ratios, insufficient high-coverage bases, elevated chimeric read fractions, low call rates across high-confidence sites, sex inconsistencies, and cryptic relatedness.

Additional outlier detection was performed on key sequencing and variant metrics, including indel ratio, transition-to-transversion ratio, singleton-to-transition ratio, and heterozygosity rate. These metrics were evaluated after stratification by sequencing platform and adjustment for population structure through regression on the top 20 principal components; samples exceeding outlier thresholds were removed.

Genetic ancestry assignments provided by gnomAD were incorporated directly. Ancestry inference was conducted by clustering samples based on genetic similarity using Hardy–Weinberg equilibrium–normalized principal component analysis across 30 principal components derived from high-quality variant sites. The leading 20 components were subsequently used to train a random forest classifier, which assigned ancestry labels based on posterior probability thresholds. Samples lacking an ancestry classification were excluded from downstream analyses (**Supplementary Figure 1**).

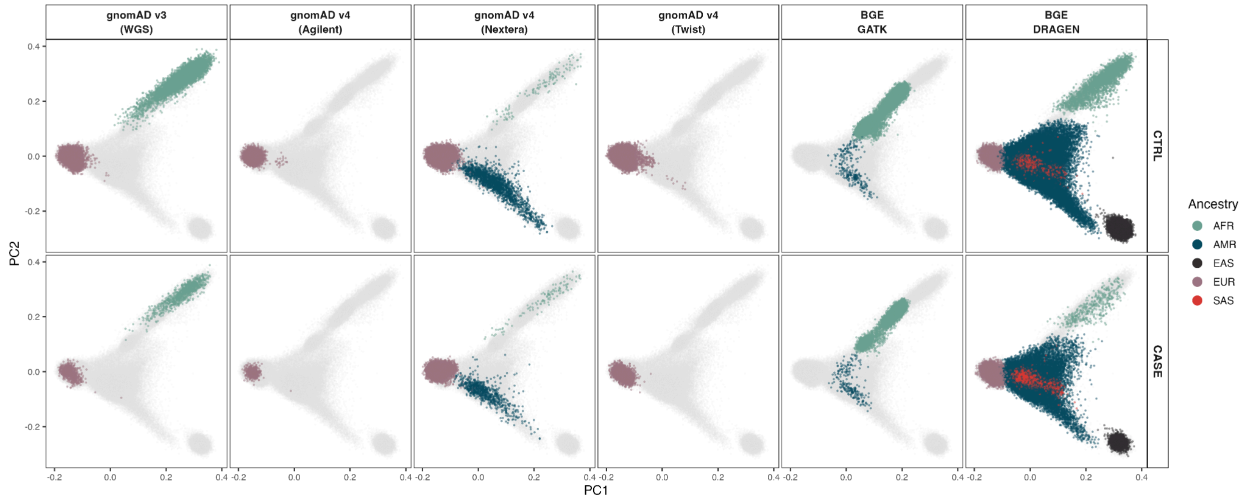


**Supplementary Figure 1. Population structure and ancestry-stratified case-control distribution across each sequencing platform.** Principal component analysis (PCA) plots illustrate the genetic diversity and phenotypic distribution specifically within each sequencing technology. The top-left panel displays all individuals projected onto the reference ancestry space (gray), colored by inferred global genetic ancestry (AFR, teal; AMR, dark blue; EAS, black; EUR, mauve; SAS, coral). The close overlap of cases and controls within each ancestry-specific cluster demonstrates that phenotypic status is not confounded by sub-population structure, supporting the validity of the stratified Cochran-Mantel-Haenszel framework used for association testing.

Additionally, for samples with assigned ancestry, quality metrics were computed using Hail's sample_qc() function. Within each cohort–ancestry stratum, outlier removal was performed using median absolute deviation (MAD) thresholds. Samples exceeding ±4 MADs from the median were removed based on transition-to-transversion ratio (Ti/Tv), counts of insertions, deletions, transitions, transversions, total SNPs, heterozygosity ratio, and indel ratio. Singleton counts were treated asymmetrically due to their lower bound at zero; samples were excluded if values were more than 8 MADs above the median. Additionally, a maximum threshold of 5000 was imposed on gnomAD v4.0 exome samples, to be consistent with subsequent analyses of gnomAD v4.0 samples (**Supplementary Figure 2**). An identical procedure was applied to gnomAD v3.0 genome samples, without the additional singleton hard filter of 5000 (**Supplementary Figure 3**).

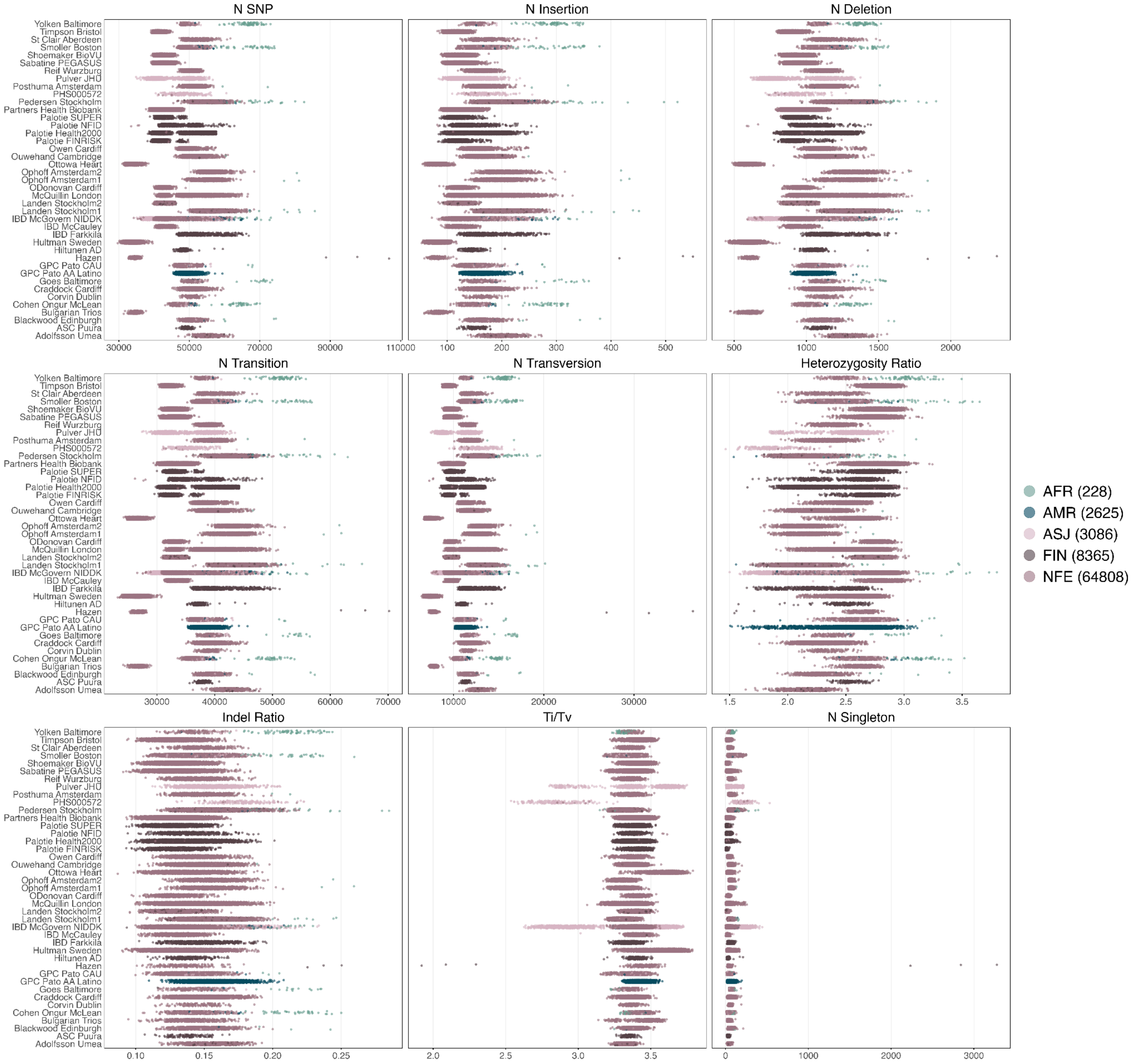


**Supplementary Figure 2. Distribution of sample quality control (QC) metrics across gnomAD v4.0 exome cohorts.** Dot plots illustrate the distribution of nine key sequencing metrics for samples passing all QC filters, stratified by cohort (y-axis) and inferred genetic ancestry (color: AFR, teal; AMR, dark blue; ASJ, light pink; FIN, dark maroon; NFE, mauve). Metrics were generated using the Hail sample_qc() function and include total counts of SNPs, insertions, deletions, transitions (Ti), transversions (Tv), and singletons, alongside calculated ratios for heterozygosity, indels, and Ti/Tv. Each point represents an individual, with the horizontal spread demonstrating natural variation across the diverse global cohorts contributing to the dataset. Distributions reflect the final sample set after outlier removal within each cohort–ancestry stratum using median absolute deviation (MAD) thresholds; standard metrics were filtered at $\pm4$ MADs, while singleton counts—which are highly sensitive to sample composition and zero-bounded—were filtered asymmetrically at $+8$ MADs above the median.

Additionally, a strict maximum threshold of 5,000 singleton counts was imposed to maintain strict conformity with downstream analysis requirements.

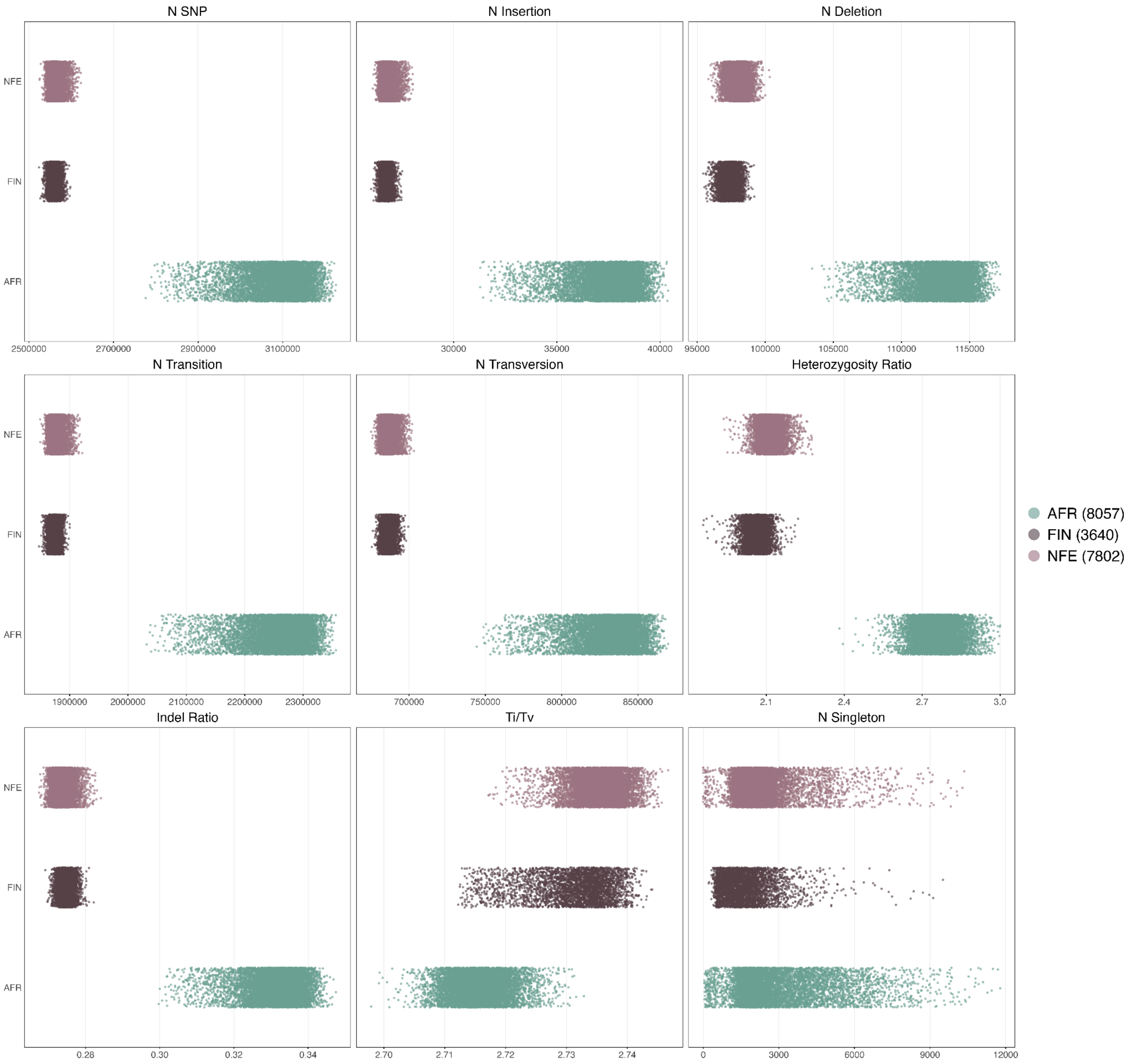


**Supplementary Figure 3. Distribution of sample quality control (QC) metrics across gnomAD v3.0 genome cohorts.** Dot plots illustrate the distribution of nine key sequencing metrics for whole-genome sequenced (WGS) samples passing all QC filters, stratified by inferred genetic ancestry on the y-axis (color: AFR, teal; FIN, dark brown; NFE, mauve). Metrics were generated using the Hail sample_qc() function and include total counts of SNPs, insertions, deletions, transitions (Ti), transversions (Tv), and singletons, alongside calculated ratios for heterozygosity, indels, and Ti/Tv. Each point represents an individual sample, with the horizontal spread demonstrating natural variation across population strata. Distributions reflect the final sample set after outlier removal using median absolute deviation (MAD) thresholds within each ancestry stratum; standard metrics were filtered at $\pm 4$ MADs, while singleton counts were filtered asymmetrically at $+8$ MADs above the median. An identical procedure to the exome pipeline was applied here, but without the additional upper hard filter of 5,000 on singletons.

#### Sample Quality Control and Ancestry Inference for BGE data

Analyses of the BGE-GATK dataset were limited to the exome component and to samples meeting stringent quality criteria, including hard filters, genetic ancestry assignment, and statistical outlier removal. Samples were first excluded if chimeric read fractions or estimated contamination exceeded 5% when the data were available. Genetic ancestry was inferred using principal component analysis coupled with supervised classification. Because of the distinctive population structure represented in the BGE cohort, a custom ancestry reference panel was assembled. This panel integrated samples from the Human Genome Diversity Project (HGDP), the 1000 Genomes Project Phase 3, the African Wits-INDEPTH Partnership for Genomic Studies (AWI-Gen), and the African Genome Variation Project (AGVP). BGE samples were merged with the reference panel and filtered to variants with call rates above 99% and minor allele frequency exceeding 0.1%. Linkage disequilibrium pruning was performed to retain approximately independent variants, after which the top 10 principal components were computed using Hail's hwe_normalized_pca().

For samples with assigned ancestry, quality metrics were computed using Hail's sample_qc() function. Within each cohort–ancestry stratum, outlier removal was performed using median absolute deviation (MAD) thresholds. Samples exceeding ±4 MADs from the median were removed based on transition-to-transversion ratio (Ti/Tv), counts of insertions, deletions, transitions, transversions, total SNPs, heterozygosity ratio, and indel ratio. Singleton counts were treated asymmetrically due to their lower bound at zero; samples were excluded if values were more than 8 MADs above the median (**Supplementary Figure 4**). This is due to singletons being highly sensitive to the sample composition. We applied similar sample quality control for the BGE-DRAGEN dataset (**Supplementary Figure 5**).

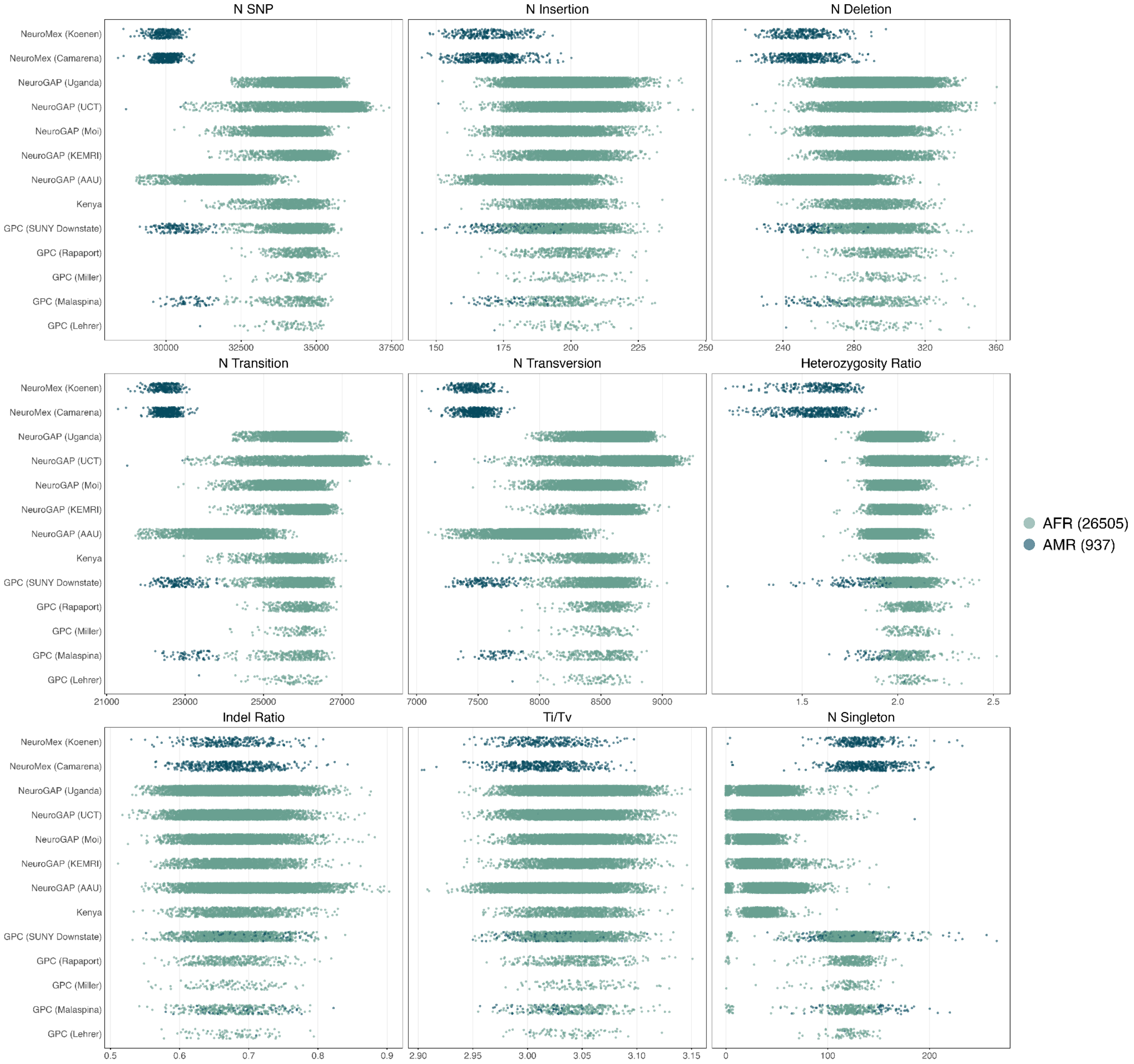


**Supplementary Figure 4: Distribution of sample quality control (QC) metrics across BGE-GATK cohorts.** Dot plots illustrate the distribution of nine key sequencing metrics for samples passing all QC filters, stratified by cohort (y-axis) and inferred genetic ancestry (color: AFR, teal; AMR, dark blue). Metrics were generated using the Hail sample_qc() function and include total counts of SNPs, insertions, deletions, transitions (Ti), transversions (Tv), and singletons, alongside calculated ratios for heterozygosity, indels, and Ti/Tv. Each point represents an individual, with the horizontal spread demonstrating natural variation across recruitment sites in Africa and the Americas. Distributions reflect the final sample set after outlier removal using median absolute deviation (MAD) thresholds; standard metrics were filtered at $\pm 4$ MADs, while singleton counts—which are highly sensitive to sample composition and zero-bounded—were filtered asymmetrically at $+8$ MADs.

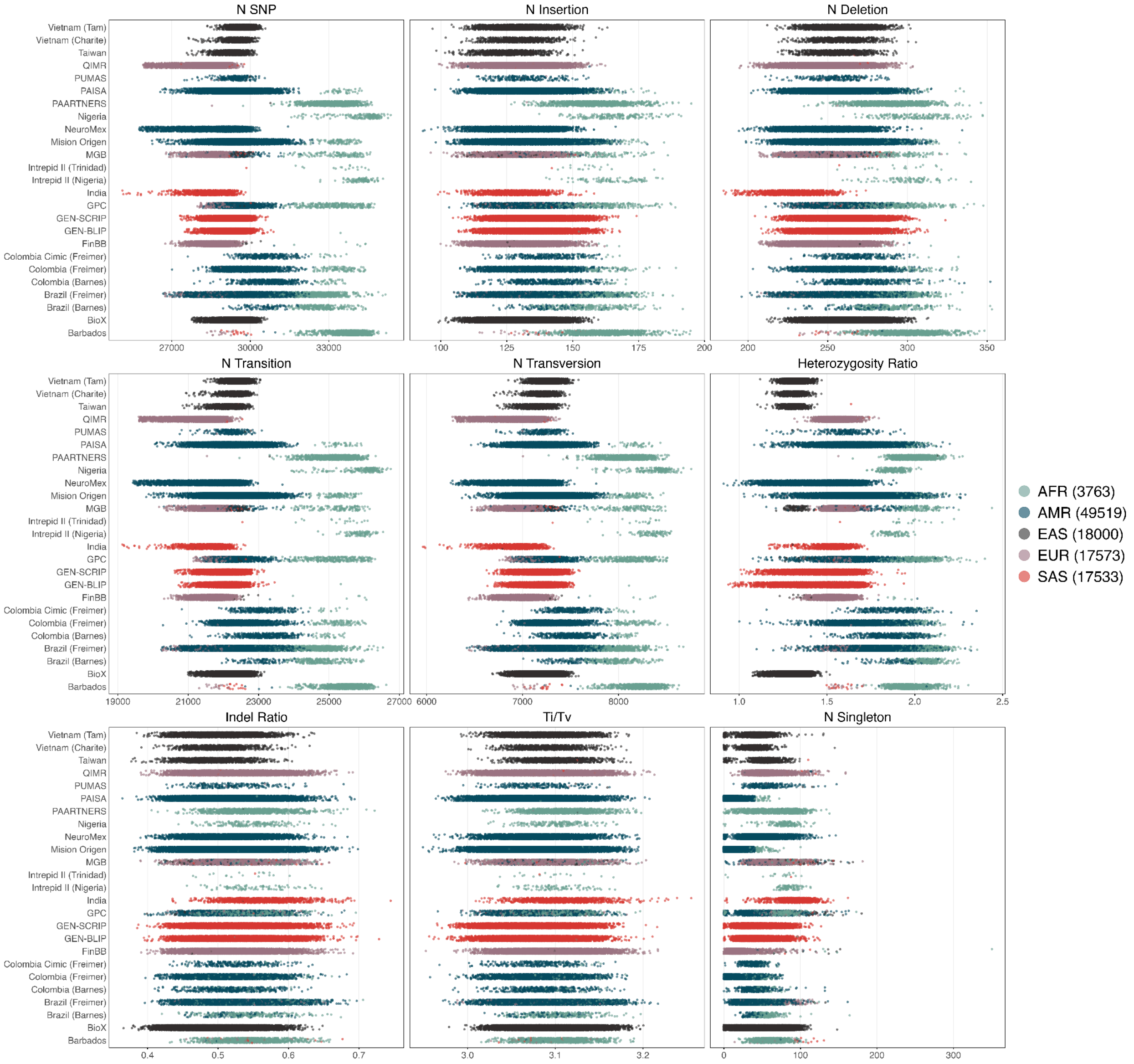


**Supplementary Figure 5: Distribution of sample quality control (QC) metrics across BGE-DRAGEN cohorts.** Dot plots illustrate the distribution of nine sequencing metrics for samples passing all QC filters within the DRAGEN processing pipeline, stratified by cohort (y-axis) and inferred global genetic ancestry (color: AFR, teal; AMR, dark blue; EAS, dark grey; EUR, mauve; SAS, coral). All sample-level metrics were computed using Hail's sample_qc() function and include total counts of SNPs, insertions, deletions, transitions (Ti), transversions (Tv), and singletons, alongside ratios for heterozygosity (het/hom-var), indels (insertion/deletion), and Ti/Tv. Each point represents an individual sample, with cohorts representing global recruitment sites across Africa, the Americas, South Asia, and East Asia. Visualized distributions represent the final high-quality sample set after outlier removal; samples were excluded if they exceeded 4 median absolute deviations (MADs) from the cohort-ancestry median for standard metrics, while singleton counts were filtered asymmetrically at +8 MADs due to their sensitivity to sample composition and zero-bound distribution.

Sex concordance checks were conducted to identify mismatches between genetic and reported sex. Biallelic variants on chromosome X were filtered to sites with call rates >99% and allele frequency >0.1%, followed by linkage disequilibrium pruning. Hail's impute_sex() function was then applied to estimate inbreeding coefficients (F-statistics) using variants outside pseudoautosomal regions. Samples with F-statistic <0.6 were classified as female and those with F-statistic >0.6 as male. Individuals with discordance between inferred genetic sex and reported gender were removed.

For both the BGE-GATK and BGE-DRAGEN data, we calculated the number of chrY genotype calls for each samples as a proxy for genetic sex. Samples with discordant genetic sex and reported gender were moved. For BGE-GATK, samples with < 400 calls were labelled female and samples with > 500 were labelled as male (**Supplementary Figure 6**).

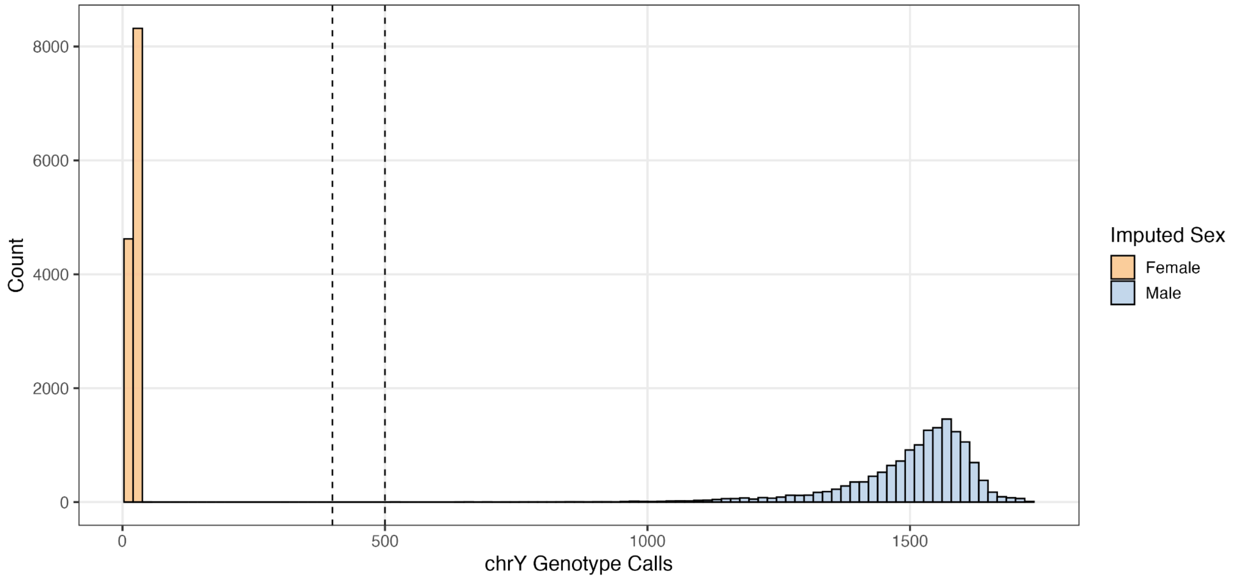


**Supplementary Figure 6: Genetic sex imputation based on chrY genotype calls in BGE-GATK.** Histogram illustrating the distribution of non-missing genotype calls on chromosome Y (chrY) used to infer genetic sex. Samples were classified as female (orange) if they possessed $< 400$ chrY genotype calls and as male (blue) if they possessed $> 500$ calls, with thresholds indicated by vertical dashed lines. This metric serves as a proxy for genetic sex; samples exhibiting discordance between inferred genetic sex and self-reported gender were excluded to ensure sample integrity. The specific call-count distribution is a function of the BGE-GATK sequencing capture efficiency and the variant calling pipeline employed. The clear bimodal separation indicates robust discrimination between sex groups, with a minimal number of samples falling within the indeterminate range (400–500 calls).

For BGE-DRAGEN, samples with < 4500 calls were labelled female and samples with > 4500 were labelled as male (**Supplementary Figure 7**).

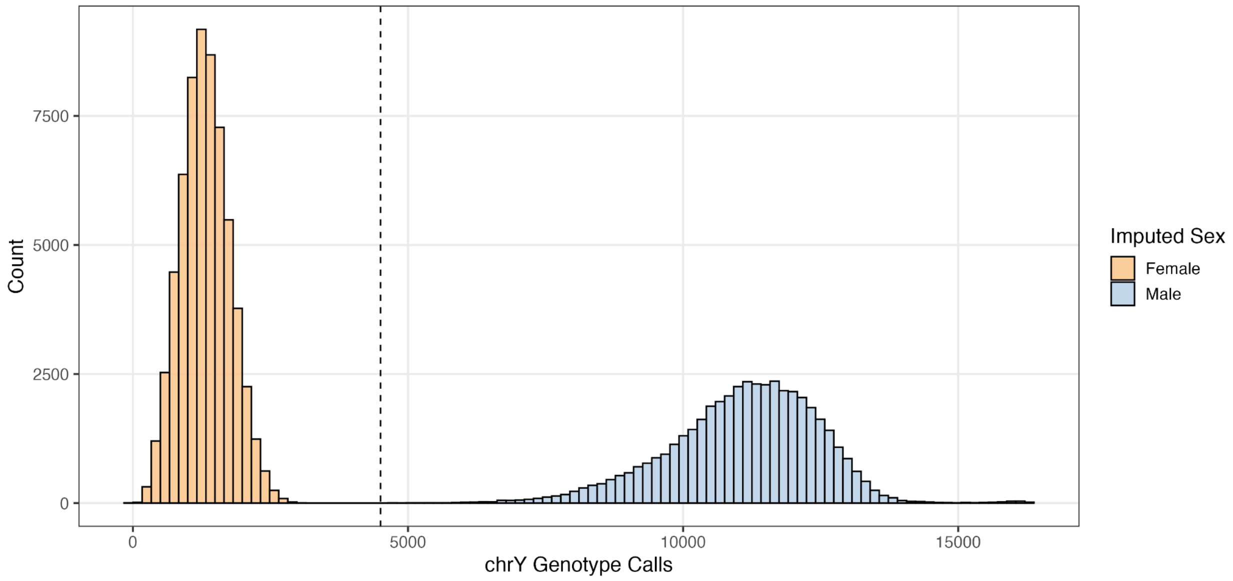


**Supplementary Figure 7: Genetic sex imputation based on chrY genotype calls in BGE-DRAGEN.** Histogram illustrating the distribution of non-missing genotype calls on chromosome Y (chrY) used to infer genetic sex for the DRAGEN-processed sub-cohort. Due to the higher sensitivity and variant calling properties of the DRAGEN pipeline, a single threshold of 4,500 calls (indicated by the vertical dashed line) was utilized; samples with fewer than 4,500 calls were classified as female (orange), while those with greater than 4,500 calls were classified as male (blue). This metric was used to identify and resolve instances of discordance between inferred genetic sex and reported sex. The wide separation between the female and male peaks indicates high confidence in sex assignment, while the distinct rightward shift in total genotype calls compared to the BGE-GATK dataset (**Supplementary Figure 6**) highlights the necessity of pipeline-specific calibration for sex imputation.

Following callset-specific quality control, we merged all passing samples from the different cohorts into a single Hail MatrixTable.

#### Relatedness

Following callset-specific quality control, we merged all passing samples from the different cohorts into a single Hail MatrixTable. To assess relatedness, we focused on variants with a callrate >0.99 and a minor allele frequency greater than 1%. On the combined dataset, we filtered to LD-pruned variants part of the gnomAD v4.0 PCA loadings. We tested both PC-Relate and KING for calculating relatedness and used independently sequenced duplicated samples as a positive control. We moved forward with KING given that PC-Relate provided unstable estimates of kinship (outside of expected ranges) with duplicated samples. Individuals with genetic relatedness >0.125 were removed (**Supplementary Figure 8**).

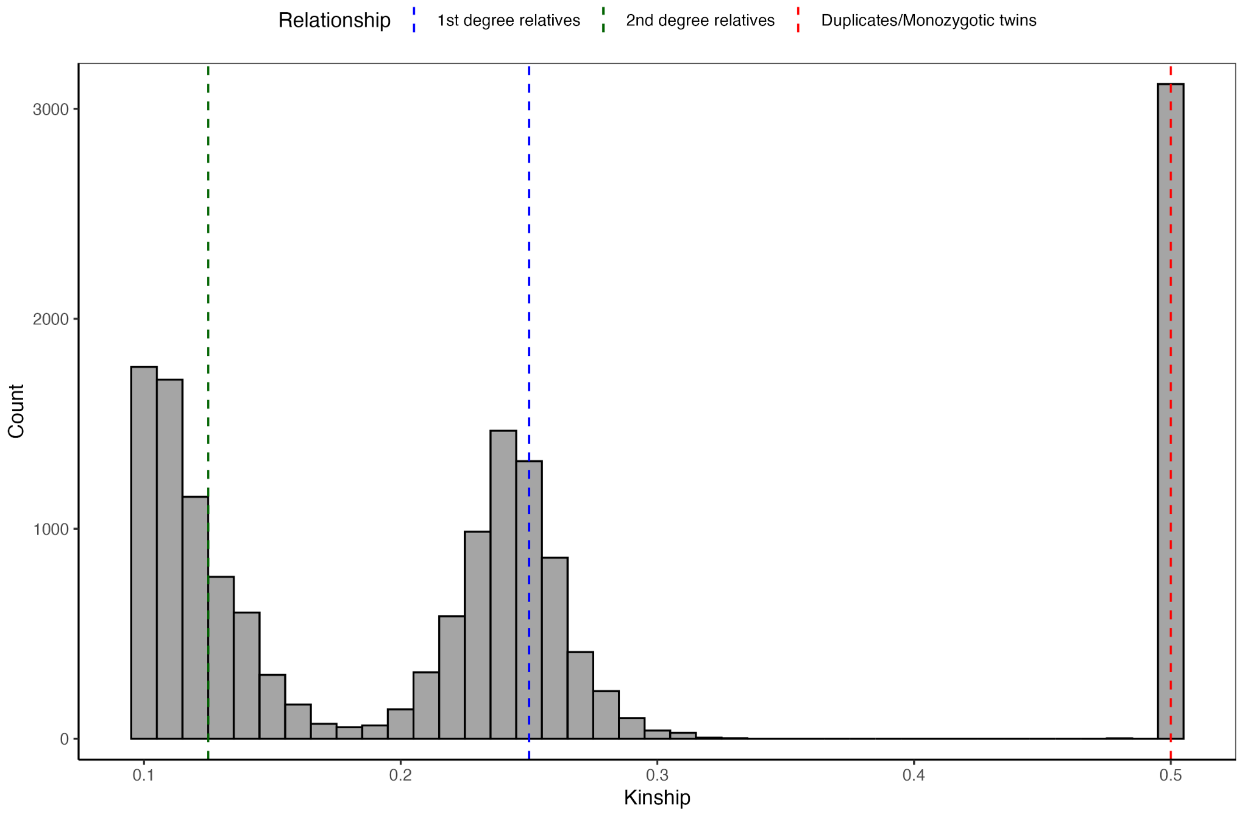


**Supplementary Figure 8. Distribution of kinship coefficients across the callset.** Histogram displaying the distribution of kinship coefficients calculated using KING (x-axis) for all pairwise sample comparisons. The bimodal distribution shows clear clusters corresponding to different degrees of relatedness. Vertical dashed lines indicate theoretical thresholds for expected relationships: 2nd degree relatives ($\geq 0.125$, green), 1st degree relatives ($\geq 0.25$, blue), and duplicates or monozygotic twins ($\geq 0.5$, red). Independently sequenced duplicated samples were used as a positive control, correctly clustering at the $\geq 0.5$ threshold, thus confirming the robustness of the KING algorithm for this dataset. Individuals with genetic relatedness exceeding the 2nd degree threshold of 0.125 were removed to maintain sample independence; during the maximally independent set procedure, case samples were preferentially retained over control samples to maximize phenotypic power. Additionally, aberrant samples with more than 10 detected relations were excluded as likely low-quality or contaminated samples.

#### Annotating variants

To annotate variants, we used Ensembl's Variant Effect Predictor (VEP) v95. All annotations are based on the Ensembl canonical transcript.

##### Protein-truncating variants (PTVs)

Variants annotated as transcript ablation, splice acceptor, splice donor, stop-gain, or frameshift were classified as protein-truncating variants (PTVs). High-confidence loss-of-function variants were identified using the LOFTEE v1 framework with minor modifications.

After restricting to LOFTEE-designated high-confidence PTVs, additional filters were applied to remove variants flagged as occurring in single-exon genes or lacking exon annotations due to intronic overlap. Furthermore, to address annotation gaps in GRCh38 LOFTEE v1 related to GERP scores, we excluded variants violating the 50 base pair truncation rule that also had GERP distance values below zero. These variants are typically located near transcript termini, are unlikely to result in true truncation, and exhibit low evolutionary conservation.

#### Defining damaging missense

To define damaging missense variants, we leveraged a conservative ensemble approach which included MPC v2, AlphaMissense, and MisFit-S. To integrate these methods, we developed a mean-missense Z-score approach (MeanMis). Briefly, each missense score is transformed across the exome into a rank. Subsequently, the ranks are converted into Z-scores, which accounts for the varying lengths of scores within MPC v2, AlphaMissense and MisFit-S. We took the mean rank Z-score and defined damaging as >96%. The threshold was defined empirically by assessing where enrichment stagnates across the exome-wide BipEx data (**Supplementary Figure 9**).

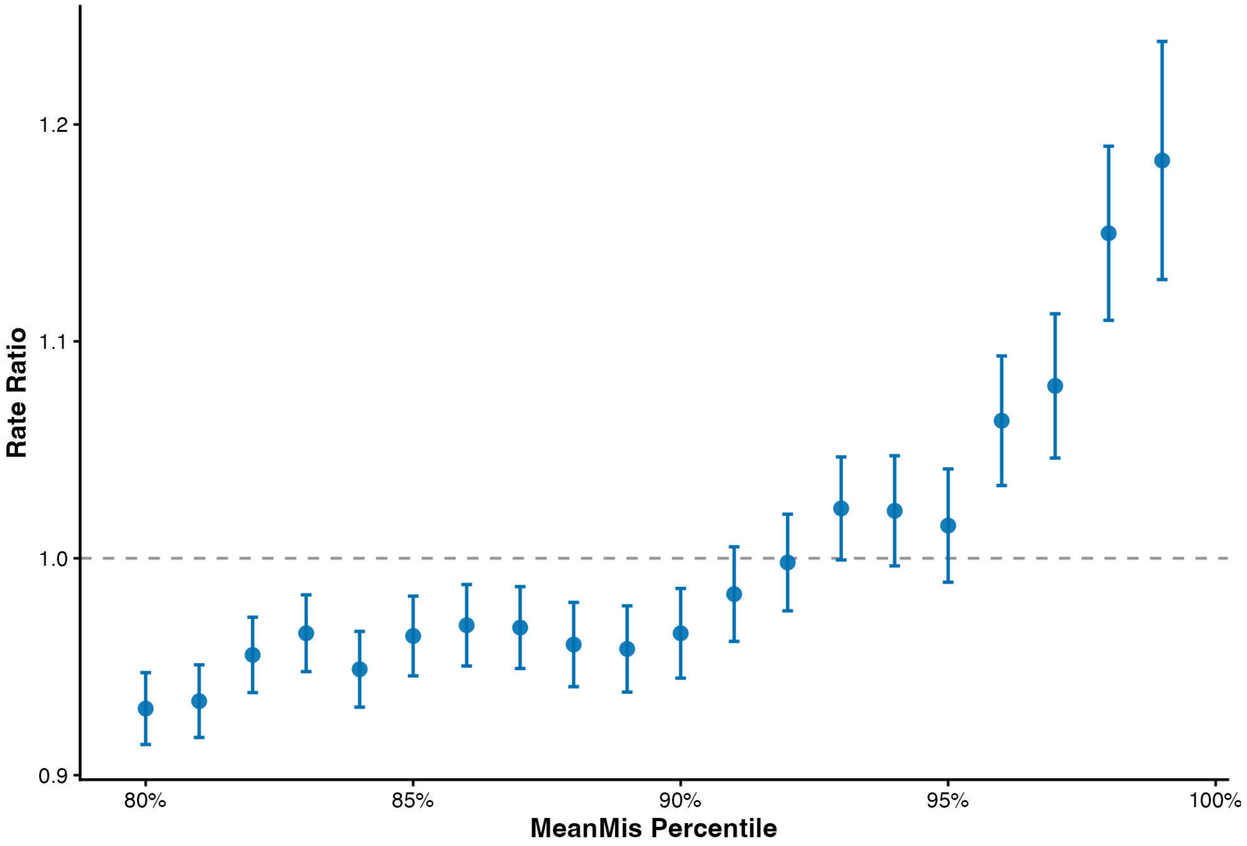


**Supplementary Figure 9. Empirical determination of the MeanMis threshold for damaging missense variants.** Plot illustrating the rate ratio (y-axis) of bipolar case versus control carrier status within constrained genes, evaluated across varying MeanMis percentile bins (x-axis). Points represent the case-control rate ratio for each MeanMis percentile bin, with vertical bars indicating standard error. The horizontal dashed line denotes the null expectation (Rate Ratio = 1.0). The threshold defining "damaging" variants was empirically established at >96%, corresponding to the critical point in the BipEx dataset where variant enrichment clearly diverges from the null and sharply increases, in contrast to the stagnant enrichment observed at lower percentiles.

#### Gene set burden testing

We performed gene set burden analyses using sample level rare variant burden counts aggregated by gene set and variant consequence class. Variant classes included protein truncating variants, damaging missense variants, a combined class of protein truncating and damaging missense variants, and synonymous variants. Analyses were conducted separately for each variant class and gene set.

Burden tests were performed within each major ancestry group using logistic regression, modeling bipolar disorder case status as the outcome and the gene set burden as the primary predictor. All models included sex, the first ten ancestry principal components, and sequencing platform as covariates. Analyses were restricted to ancestry strata with at least ten cases and ten controls, and gene set tests required a minimum of five carriers within each ancestry stratum.

Effect estimates from ancestry specific models were combined using fixed effect inverse variance weighted meta analysis on the log odds ratio scale. For each gene set and variant class, we report the meta analyzed odds ratio, 95% confidence interval, and two sided P value. Between ancestry heterogeneity was assessed using Cochran’s Q statistic and summarized with the $I^2$ metric.

All analyses were performed using R, with logistic regression implemented via the glm function and meta analysis conducted using custom inverse variance weighting procedures.

#### Rare variant association testing

For the rare variant association testing, we compared both the Fisher's Exact Test and CMH test. We found that Fisher's Exact was highly inflated due to extensive strata from multiple sequencing platforms and ancestry relative to the CMH test (**Supplementary Figure 10**). We restricted to variants with a minor allele frequency <=0.01%. For the CMH test, we removed ancestry-by-cohort groups that had fewer than 50 cases, fewer than 5 controls, or fewer than 200 total samples due to small sample size and the potential to bias the test.

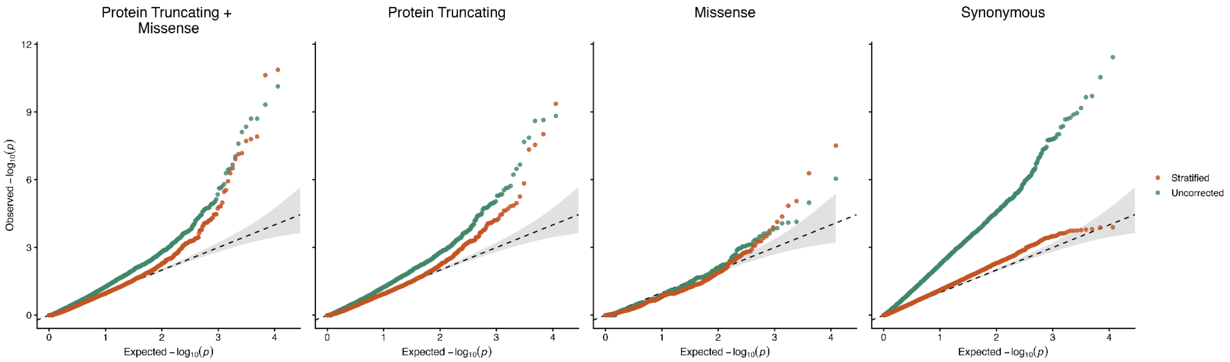


**Supplementary Figure 10: Q-Q plots for rare variant gene-based burden comparing Fisher's Exact Test and CMH Test.** Distributions of observed vs. expected P-values are shown for four functional categories (Protein Truncating + Damaging Missense, Protein Truncating, Damaging Missense, and Synonymous). To account for potential confounders, P-values were derived from a Cochran-Mantel-Haenszel (CMH) framework, stratifying by sequencing platform and inferred genetic ancestry.

##### Fisher comparison

To minimize the risk of false-positive associations stemming from technical batch effects and population stratification, we employed a stratified CMH testing framework. Unlike a standard Fisher's Exact test—which assumes a homogeneous population, the CMH test allows for the control of covariates by partitioning the data into strata based on sequencing platform and inferred ancestry.

As shown in **Supplementary Figure 10**, the CMH framework maintains strong calibration in the synonymous 'negative control' category. In contrast, a Fisher's Exact approach results in substantial synonymous inflation (3.01), likely driven by differences in sequencing depth and ancestral allele frequency shifts that are not properly accounted for in an unstratified model.

#### Cross-disorder rare-variant overlap analysis.

We defined the BD gene universe as protein-coding genes reaching Bonferroni significance in the combined PTV+missense rare-variant association analysis ($q < 0.05$). External gene sets were curated as follows: SCZ genes were defined as the FDR 5% significant genes ranked by rare-variant case-control association; ASD genes were defined as those with FDR < 0.001 in published exome association results; and DD genes were defined as genes annotated as NDD373-positive in exome TADA analyses from Fu et al. 2022. Gene symbols were standardized and matched across datasets using upper-case HGNC symbols. Overlap was visualized using a dot-matrix representation anchored to BD-significant genes, with connectors indicating genes shared across ≥2 external phenotypes.

#### Partitioned heritability

Partitioned heritability was estimated using stratified LD score regression (LDSC). We constructed a custom genomic annotation consisting of the 13 Bonferroni-significant genes identified in the gene-based rare variant analysis, defined as each gene body with ±100 kb flanking regions to capture local regulatory variation. This annotation comprised approximately 0.14% of all SNPs.

Stratified LDSC was applied to genome-wide association summary statistics for psychiatric and non-psychiatric traits using the baseline LD v2.2 model as a covariate set. Enrichment was calculated as the proportion of SNP heritability explained by the annotation divided by the proportion of SNPs in the annotation. Standard errors and *P* values were estimated using the block jackknife procedure implemented in LDSC. All analyses were restricted to well-imputed HapMap 3 SNPs and used European ancestry LD scores. Bonferroni correction was performed adjusting for the number of tests.

Traits analyzed included schizophrenia, bipolar disorder, autism spectrum disorder, major depressive disorder, attention-deficit/hyperactivity disorder, height, and coronary artery disease.  Enrichment estimates were interpreted relative to the null expectation of no enrichment (enrichment = 1).

#### Pathway enrichment of associated BD genes

Genes were selected from BipEx gene-level burden analyses based on a nominal association threshold of $P < 0.01$. To reduce potential bias from reverse causation or control-specific comorbidities (e.g., cardiovascular or lipid phenotypes), only genes exhibiting increased burden in cases (odds ratio > 1) were retained for downstream analyses. This approach was designed to increase sensitivity for detecting convergent biological pathways while maintaining interpretability of effect direction.

Pathway enrichment analysis was performed using ToppGene Suite (ToppFun). The input gene list was tested for enrichment across Gene Ontology categories (Biological Process and Cellular Component), curated disease annotations (e.g., DisGeNET, GWAS Catalog), and literature-derived gene sets (PubMed). Enrichment p-values were calculated using hypergeometric tests comparing the overlap between the input gene list and each annotation category relative to the genomic background.

Multiple testing correction was applied using Bonferroni correction as well as false discovery rate procedures (Benjamini-Hochberg and Benjamini-Yekutieli). Pathways reported in the main text were required to pass Bonferroni correction (pBonf < 0.05). For each enriched pathway, both the number of genes overlapping the query list and the total number of genes annotated to the pathway in the genome were recorded.

#### Transcriptomic analyses of BipEx genes

##### Gene set definition

The genes included in the expression analyses correspond to the BipEx-associated genes identified in the rare variant association analyses. Gene symbols were standardized to HGNC nomenclature and converted to uppercase prior to analysis. The final gene set consisted of thirteen genes: *ATP2B2*, *SP4*, *RB1CC1*, *SHANK1*, *DOP1A*, *KDM5B*, *ATP9A*, *CUL1*, *EIF4E2*, *EIF4A2*, *HECTD2*, *TERF2*, and *AKAP11*. These genes were used as the input set for all tissue, cell-type, and developmental expression analyses.

##### GTEx Bulk tissue expression analysis

Bulk tissue expression patterns were evaluated using transcriptomic data from the GTEx v8 project through the FUMA gene2func framework. Average normalized expression values across tissues were extracted and organized into gene-by-tissue matrices using standardized gene symbols. When duplicate gene entries were present, expression values were collapsed by averaging across entries. For visualization, expression values were limited to a range of −2 to 2. Hierarchical clustering of genes and tissues was performed using Euclidean distance and complete linkage, and heatmaps were generated using the pheatmap package in R. To assess tissue enrichment, we used the differential expression enrichment framework implemented in

FUMA gene2func. This analysis evaluates whether the input gene set is overrepresented among genes that are significantly up- or down-regulated in specific tissues relative to others.

#### Single-Cell Expression Analysis of BipEx Genes Across Brain Cell Classes

Single-cell gene expression summaries were obtained from the CELLxGENE brain dataset export, which provides gene-by–cell-type aggregated statistics including total cell counts, mean normalized expression, and the number of cells expressing each gene. Analyses were restricted to entries labeled as brain tissue, and rows labeled “aggregated” were excluded to avoid duplication of whole-brain summaries. We focused on 13 genes identified in the BipEx analysis: ATP2B2, SP4, RB1CC1, SHANK1, DOP1A, KDM5B, ATP9A, CUL1, EIF4E2, EIF4A2, HECTD2, TERF2, and AKAP11.

Cell-type annotations were curated prior to analysis to remove non-brain and clearly irrelevant categories using pattern-based filtering. These included lung epithelial populations, hematopoietic and erythroid lineages, immune cell subsets (e.g., T cells, B cells, dendritic cells, monocytes), retinal and eye-specific cell types, mesenchymal and fibroblast populations, and neoplastic or ontology artifact labels. Macrophage annotations were retained only when explicitly labeled as central nervous system–associated (e.g., central nervous system macrophage, perivascular macrophage, meningeal macrophage, choroid plexus macrophage), while generic macrophage labels were excluded. No minimum cell-count threshold was applied prior to collapsing cell types in order to preserve high-resolution subtype information.

Fine-grained annotations were collapsed into canonical brain cell classes using rule-based pattern matching. Collapsed categories included excitatory neurons (including layer-labeled glutamatergic and pyramidal subtypes), inhibitory neurons (GABAergic and interneuron subtypes), medium spiny neurons, monoaminergic neurons (dopaminergic, serotonergic, noradrenergic), Purkinje cells, granule neurons, neural progenitors (including radial glial and precursor states), astrocytes, oligodendrocytes, oligodendrocyte precursor cells, microglia and other CNS-associated macrophages, endothelial cells, pericyte and mural cell populations, ependymal cells, choroid plexus cells, meningeal and leptomeningeal cells, and neurons of unspecified subtype. Generic labels were not reassigned to specific subclasses to avoid biologically unsupported collapsing.

For each collapsed cell class and gene, aggregated metrics were computed across contributing subtypes. Total cells were calculated as the sum of cell counts, and expressing cells as the sum of the number of cells expressing the gene. Mean expression within each collapsed class was computed as a cell-count–weighted mean of subtype-level expression values. Percent expressing was defined as the proportion of expressing cells relative to total cells within the collapsed class. Aggregation was performed prior to applying a minimum cell-count threshold; subsequently, collapsed classes with fewer than 200 total cells were excluded. Missing values were handled conservatively such that if all contributing subtype values were missing, the aggregated metric remained missing, and no values were artificially imputed.

Absolute expression patterns were visualized using a dot plot in which dot size represents the percentage of cells expressing a given gene and color represents the log-transformed weighted mean expression (log1p), used to compress dynamic range for visualization. This transformation was applied solely for visualization.

Relative enrichment across cell classes was visualized using a supplementary heatmap constructed from log-transformed mean expression values. For each gene, expression values across collapsed cell classes were standardized to z-scores using NA-aware means and standard deviations, preserving missing entries rather than imputing zeros. Genes and cell classes with excessive missingness (greater than 35%) were excluded from the heatmap. Genes were hierarchically clustered using Euclidean distance and complete linkage to identify similarities in relative cell-type enrichment patterns, while cell classes were displayed in biologically interpretable order. In addition, k-means clustering was performed on gene-level z-scored profiles to explore potential grouping of genes with similar cross–cell-type expression patterns.

A comprehensive quality control report was generated automatically, including row counts after each filtering step, distributions of collapsed cell classes, aggregated total cell counts per class, missingness fractions per gene and per class, summary statistics for percent expressing and log-transformed expression, and complete session information. All analyses were performed in R (version 4.5.2) using data.table, ggplot2, and pheatmap.

#### Brainspan developmental expression analysis

Developmental expression patterns were assessed using transcriptomic data from the BrainSpan project through the FUMA gene2func resource. The dataset includes normalized gene expression measurements spanning prenatal development through adulthood. Expression values were organized into gene-by-age matrices using standardized gene symbols. For visualization, values were constrained to a range of −2 to 2. Genes were clustered using hierarchical clustering with Euclidean distance and complete linkage, while developmental stages were maintained in chronological order to preserve temporal structure. Developmental enrichment was evaluated using the differential expression enrichment analysis implemented in FUMA gene2func, which tests whether the input gene set is overrepresented among genes differentially expressed across developmental stages.

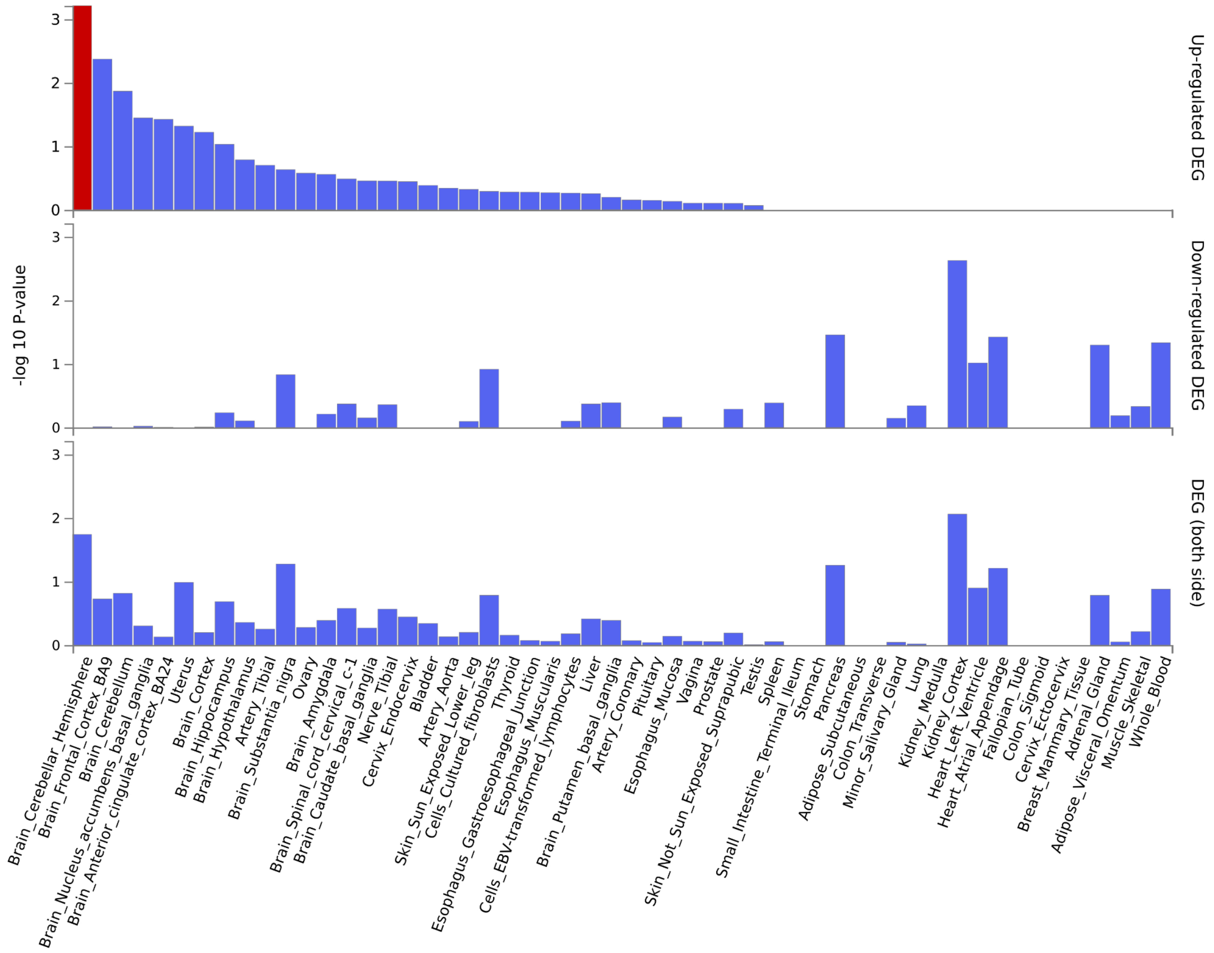


**Supplementary Figure 11. GTEx v8 tissue enrichment.** Enrichment of BipEx genes among tissue-specific DEGs across GTEx v8 tissues. Results are shown separately for up-regulated DEGs (top), down-regulated DEGs (middle), and combined (both directions; bottom). Tissues are ordered by decreasing significance. The red bar highlights the most significantly enriched tissue in each panel.

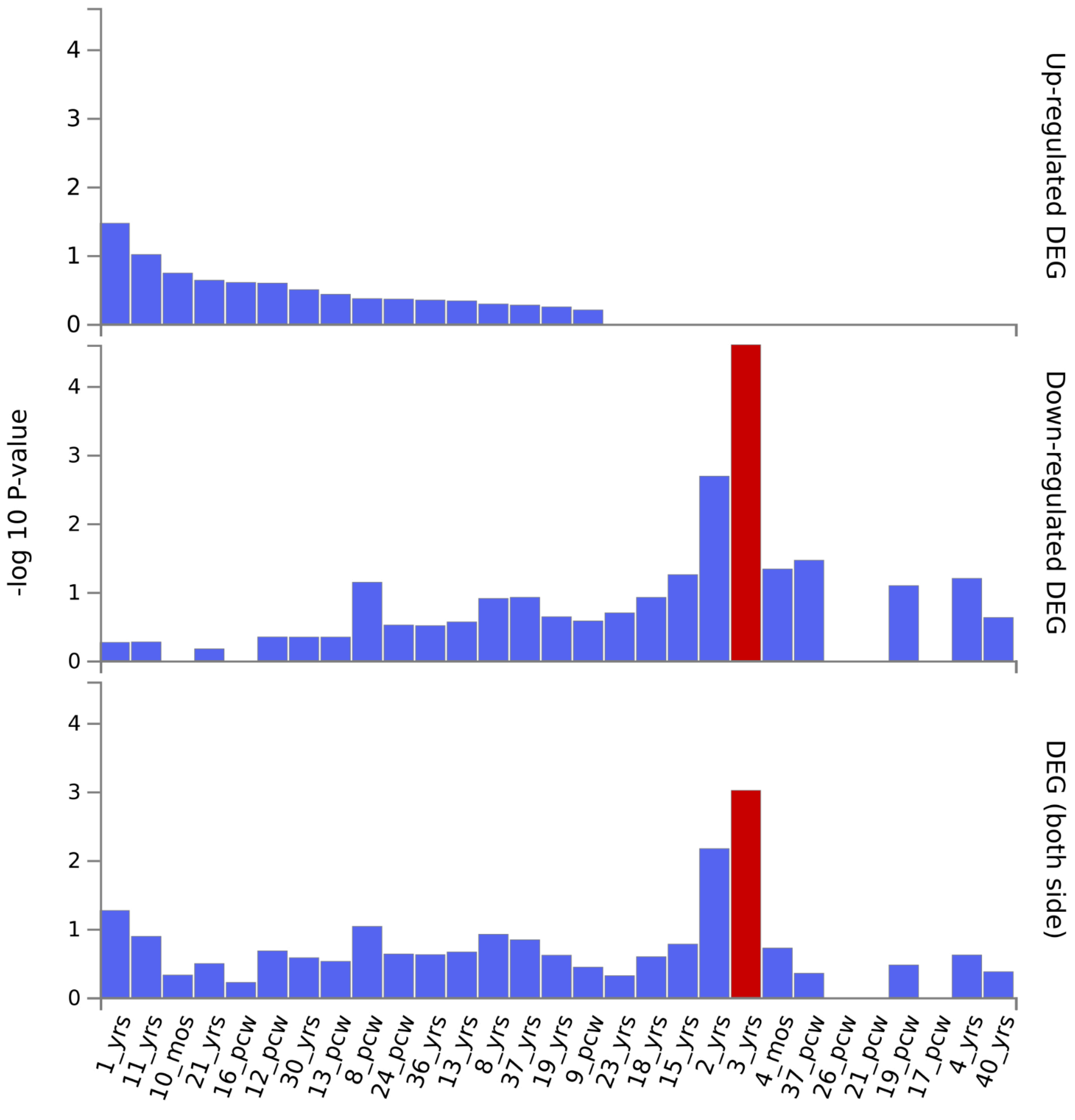


**Supplementary Figure 12. BrainSpan postnatal age enrichment.** Enrichment of BipEx genes across BrainSpan postnatal age groups. Bars represent −log10(P) values for overlap with DEGs identified at each age (years or post-conception weeks, pcw). Panels are stratified as in (a): up-regulated (top), down-regulated (middle), and combined DEGs (bottom). The red bar indicates the age group with the strongest enrichment signal.

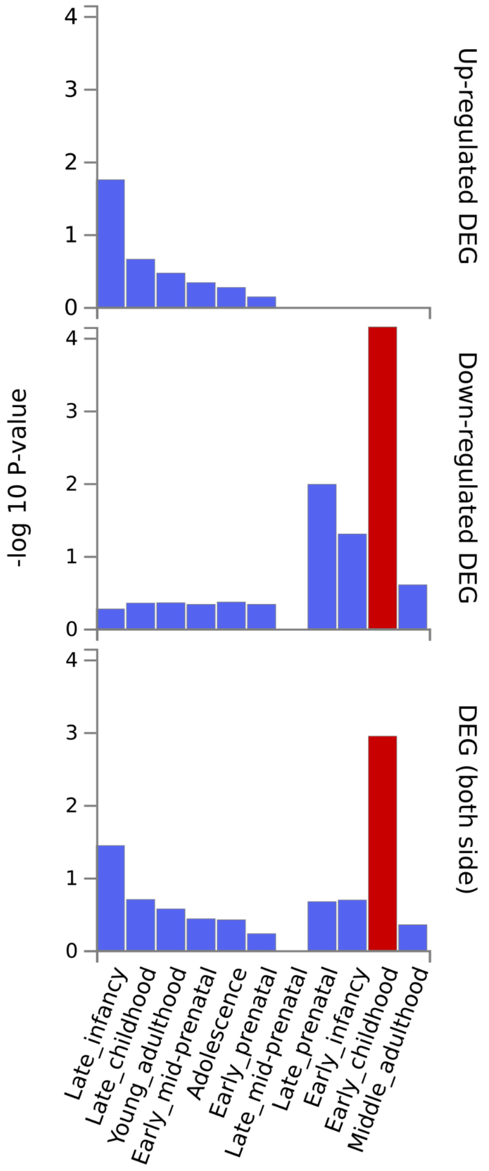


**Supplementary Figure 13. BrainSpan developmental stage enrichment.** Enrichment across broader BrainSpan developmental stages (prenatal to adulthood). As above, enrichment is shown for up-regulated, down-regulated, and combined DEGs. The red bar denotes the most significantly enriched developmental stage.

#### Identification of missense mutational hotspots using 3D protein structures

To identify spatial clustering of disease-associated missense variants, we applied the 3D Neighborhood Test (3DNT) to the top 13 BipEx genes reaching exome-wide significance. For each gene, we aggregated ultra-rare missense variants observed in the BipEx dataset.

Missense variants were mapped to canonical protein sequences and corresponding predicted three-dimensional structures using precomputed mappings from genomic coordinates to amino acid positions and AlphaFold-predicted structures. For proteins lacking structures in the AlphaFold Protein Structure Database, structures were generated using AlphaFold3. Variants that could not be confidently mapped to a protein structure were excluded.

For each residue in a protein, we defined a spherical neighborhood of radius **15 Å** centered on that residue, excluding positions with a predicted aligned error **(PAE) >15 Å** relative to the center. To identify mutational hotspots, we modeled the association between neighborhood membership and case-control status using a logistic regression framework:

$$\text{logit}(P(y_i = 1)) = \beta_0 + \beta_1 N_i + \text{logit}(b_i)$$

where $y_i$ represents the case-control status of variant $i$, $N_i$ is an indicator variable for neighborhood membership, and $b_i$ is a sample-specific covariate capturing the baseline probability of that sample's case status based on observed case/control counts in its cohort-by-ancestry strata.

To account for the large number of highly correlated tests, we employed an empirical False Discovery Rate (FDR) approach described in [Reshef et al. Nat Biotech]. The false discovery rate at a given P-value threshold is equal to the expected number of false discoveries at that threshold divided by the total number of discoveries at that threshold. We estimate the expected number of false discoveries at threshold $P_T$ by taking the average across the 1,000 null simulations of the number of neighborhoods with $P \le P_T$ in the simulation. The total number of discoveries at $P_T$ is the number of neighborhoods with $P \le P_T$ in the real data. The FDR at $P_T$ is

$$\min \left\{ \frac{\text{avg } \# \ P \leq P_T \text{ in null simulations}}{\# \ P \leq P_T \text{ in real data}} \right\}$$

where the minimum is taken over $P_{T'}>P_T$, reducing noise and ensuring that FDR is monotonic in P-value.

Neighborhoods with an FDR < 0.05 were considered significant mutational hotspots.

#### Protein-protein interaction analysis using STRING

To place hotspot-containing regions in a broader functional context, we examined known and predicted protein–protein interactions using the STRING database (v11.5). For each of the 13 BipEx genes, we retrieved first-degree interactors using default confidence thresholds. Interactors supported by experimental evidence, curated databases, or strong coexpression signals were retained.

Where available, we assessed whether interacting partners were enriched for neuropsychiatric relevance based on prior genetic association or functional annotation. STRING interaction networks were used descriptively to contextualize hotspot-containing proteins within known molecular pathways rather than as a formal enrichment analysis.

#### Structural modeling of protein-protein interactions using AlphaFold3

To assess whether mutational hotspots overlapped predicted protein–protein interaction interfaces, we used AlphaFold3 to model pairwise protein complexes between each BipEx gene product and selected high-confidence STRING interactors. For each modeled complex, we examined whether residues within significant 3DNT neighborhoods were located at or near predicted binding interfaces.

Predicted interfaces were defined based on proximity between residues across interacting proteins and qualitative inspection of AlphaFold3 confidence metrics. Overlap between mutational hotspots and predicted interaction or binding surfaces was interpreted as evidence that clustered missense variation may perturb specific molecular interactions rather than overall protein folding.

3D neighborhood test

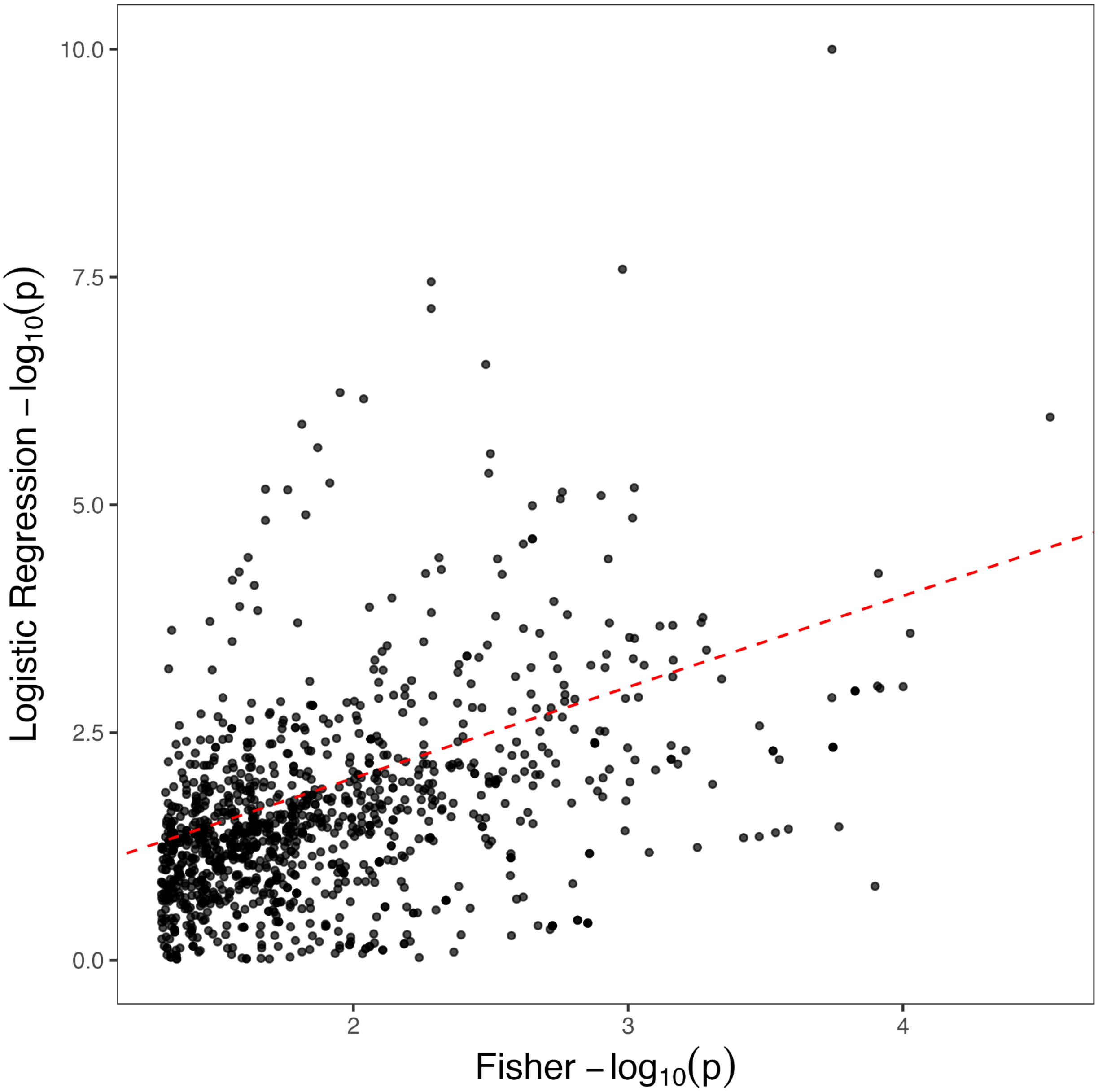


**Supplementary Figure 14. Comparison of 3D neighborhood testing under Fisher's exact test and logistic regression frameworks.** Scatter plot comparing $-\log_{10}(p\text{-values})$ derived from a standard Fisher's exact test (x-axis) and a logistic regression framework (y-axis) for protein neighborhood mutational hotspots. The logistic regression model incorporates a sample-specific covariate to account for baseline case probabilities within cohort-by-ancestry strata, whereas the Fisher's exact test does not account for this population structure. Data are filtered to points with an initial Fisher $p < 0.05$. The red dashed line indicates the $y = x$ diagonal, representing equal association strength. Points deviating from the diagonal highlight hotspots where the inclusion of the sample-specific covariate either identifies potential confounding or uncovers additional signals not captured by the univariate Fisher's exact framework.

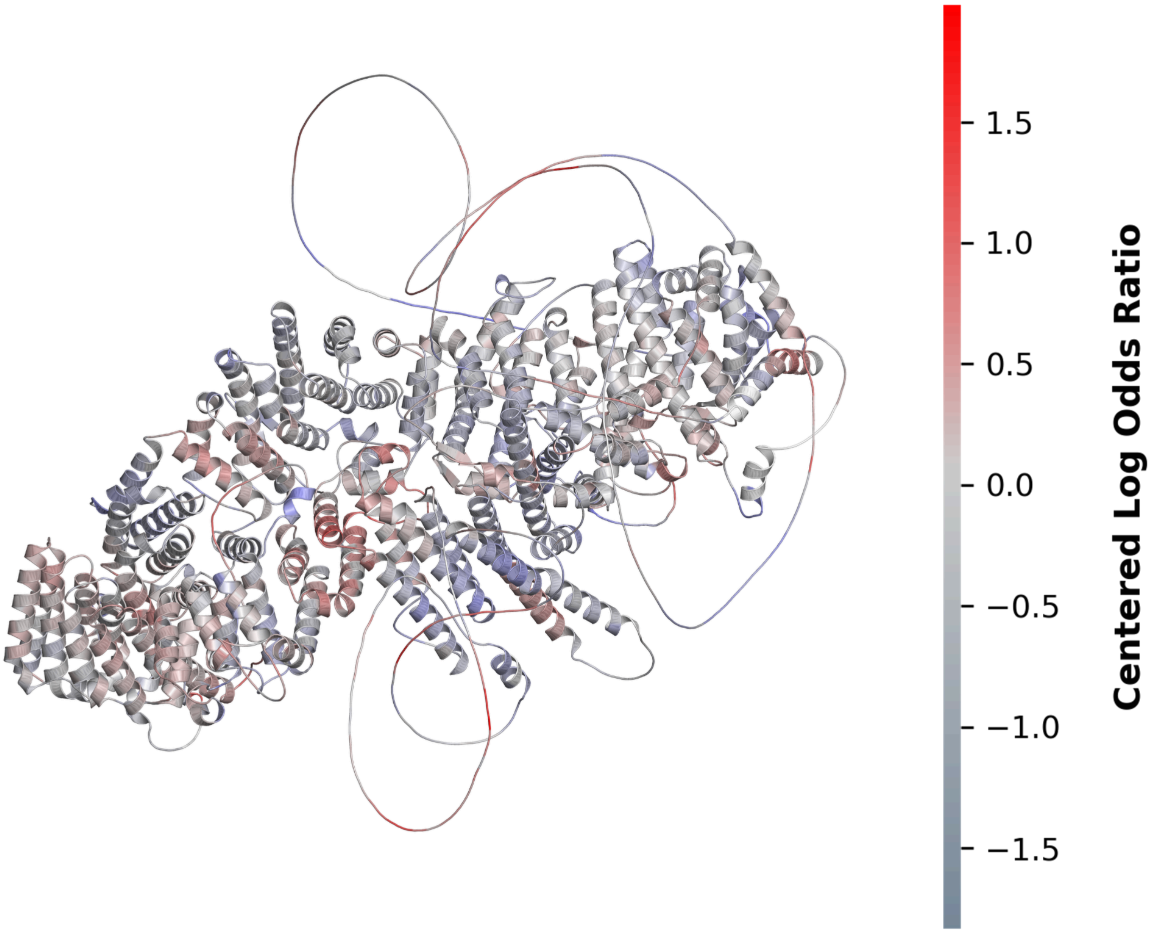


**Supplementary Figure 15. Structural context of DOP1A protein neighborhood missense associations.** Ribbon representation of the DOP1A protein structure, colored by the centered log odds ratio values derived from the logistic regression neighborhood analysis.

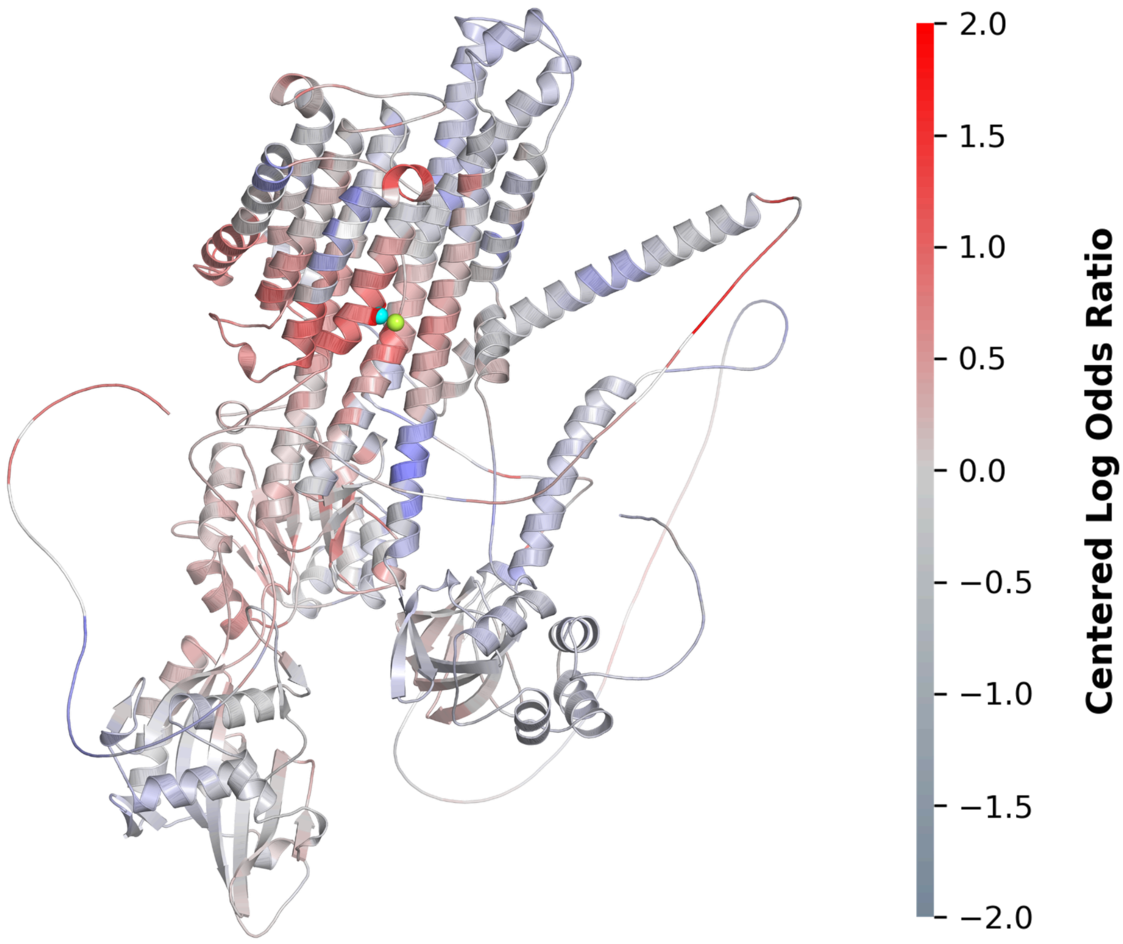


**Supplementary Figure 16. Structural context of ATP2B2 protein neighborhood missense associations.** Ribbon representation of the ATP2B2 protein structure, colored by the centered log odds ratio ratios derived from the logistic regression neighborhood analysis. Neighborhoods centered on residues **919** (cyan sphere) and **458** (limon sphere) were identified as the two strongest associations in the current analysis (centered log odds ratios of 1.819 and 1.683, respectively), though neither reached statistical significance following FDR correction (FDR = 0.325 and 0.389, respectively). Previous independent analyses in schizophrenia (Gerges et al. 2025) identified similar hotspots centered at neighboring residues 918 and 457, which constitute critical components of the core calcium-binding site. The observation that these top—albeit non-significant—associations recapitulate the location of verified risk hotspots within this conserved functional site further suggests structural consistency in mutational impact across distinct neuropsychiatric phenotypes.

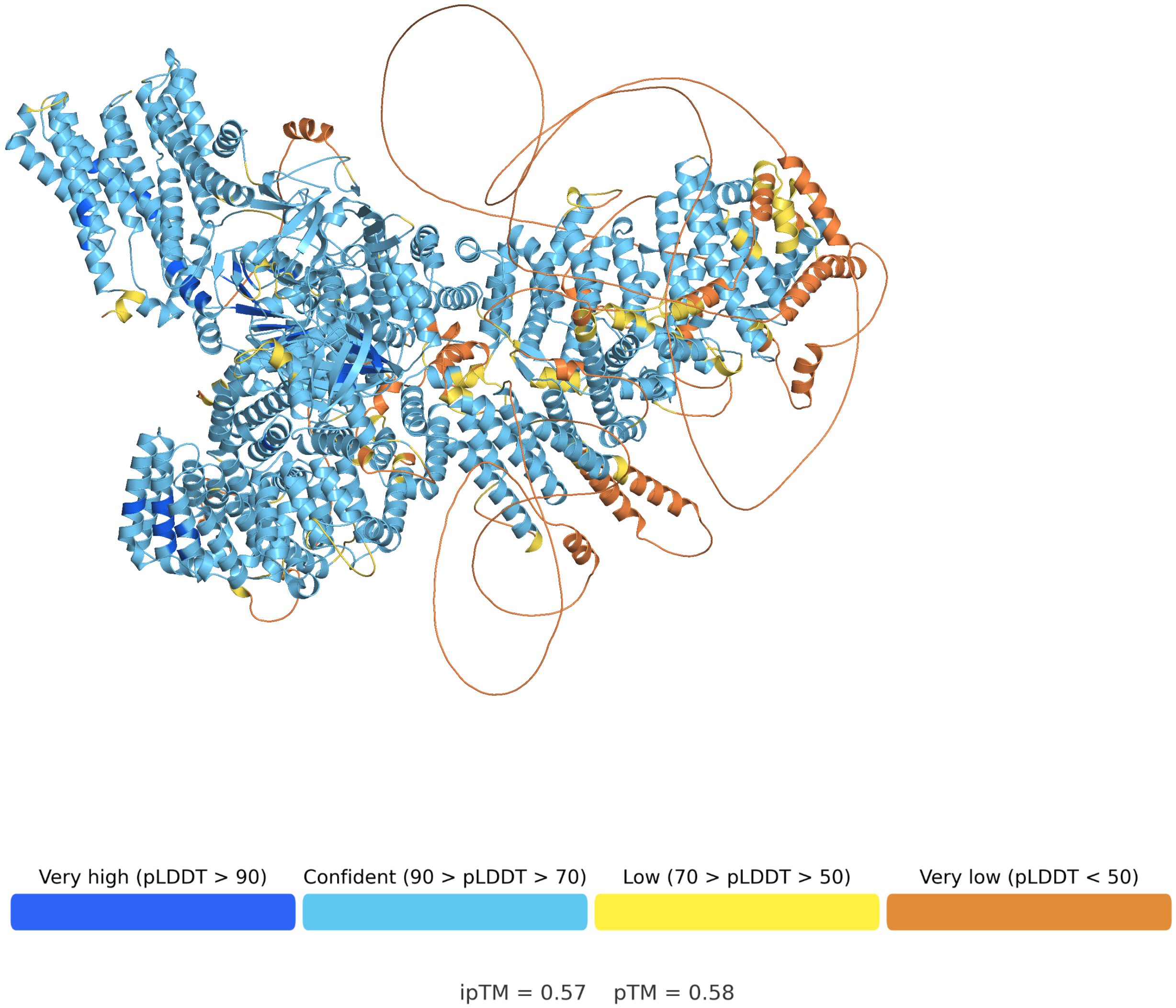


**Supplementary Figure 17. AlphaFold3 predicted structure and confidence of the DOP1A–ATP9A complex.** Ribbon representation of the predicted binary complex between DOP1A and ATP9A, colored according to the AlphaFold2 per-residue confidence metric (pLDDT). Confidence levels are categorized as very high (pLDDT > 90, blue), confident (90 > pLDDT > 70, cyan), low (70 > pLDDT > 50, yellow), and very low (pLDDT < 50, orange). The core folded domains of both proteins exhibit high to very high confidence, while the peripheral flexible loops and terminal extensions transition into lower confidence regions.
